# Substantial role of rare inherited variation in individuals with developmental disorders

**DOI:** 10.1101/2024.08.28.24312746

**Authors:** Kaitlin E. Samocha, V. Kartik Chundru, Jack M. Fu, Eugene J. Gardner, Petr Danecek, Emilie M. Wigdor, Daniel S. Malawsky, Sarah J. Lindsay, Patrick Campbell, Tarjinder Singh, Ruth Y. Eberhardt, Giuseppe Gallone, Caroline F. Wright, Hilary C. Martin, Helen V. Firth, Matthew E. Hurles

## Abstract

While the role of *de novo* and recessively-inherited coding variation in risk for rare developmental disorders (DDs) has been well established, the contribution of damaging variation dominantly-inherited from parents is less explored. Here, we investigated the contribution of rare coding variants to DDs by analyzing 13,452 individuals with DDs, 18,613 of their family members, and 3,943 controls using a combination of family-based and case/control analyses. In line with previous studies of other neuropsychiatric traits, we found a significant burden of rare (allele frequency < 1×10^-5^) predicted loss-of-function (pLoF) and damaging missense variants, the vast majority of which are inherited from apparently unaffected parents. These predominantly inherited burdens are strongest in DD-associated genes or those intolerant of pLoF variation in the general population, however we estimate that ∼10% of the excess of these variants in DD cases is found within the DD-associated genes, implying many more risk loci are yet to be identified. We found similar, but attenuated, burdens when comparing the unaffected parents of individuals with DDs to controls, indicating that parents have elevated risk of DDs due to these rare variants, which are overtransmitted to their affected children. We estimate that 6-8.5% of the population attributable risk for DDs are due to rare pLoF variants in those genes intolerant of pLoF variation in the general population. Finally, we apply a Bayesian framework to combine evidence from these analyses of rare, mostly-inherited variants with prior *de novo* mutation burden analyses to highlight an additional 25 candidate DD-associated genes for further follow up.

## Introduction

Developmental disorders (DDs) have a strong genetic component with many hundreds of genes associated with these disorders via both dominant and recessive mechanisms. However, we are able to find diagnostic genetic variants for only ∼40% of cases^1,2^, leaving over half of patients with potential genetic contributions undiagnosed. The majority of known diagnoses in cohorts such as the Deciphering Developmental Disorders (DDD) are from *de novo* variants (∼76% of diagnosed cases in DDD^2^), including *de novo* single nucleotide variants (SNVs), small insertions and deletions (indels), and structural variants (SVs) such as copy number variants. Recessive diagnoses in DDD are a distant second (∼12% of diagnosed cases) and most commonly impact families with consanguinity^2^. Large SVs, particularly at established genomic disorder loci, are well-known DD risk factors, but are often observed in unaffected family members and inherited from parents^3,4^. While the field has long appreciated incomplete penetrance of SVs^5^, there is still much to be learned about how incompletely penetrant SNVs and indels that could be inherited from unaffected parents may contribute to overall risk of DDs^6^.

We and others have previously shown that inherited common variants influence phenotypic variability^7–9^, but have not undertaken a systematic analysis of how rare, inherited variants could contribute to DDs outside of *ANKRD11*^10^. Prior studies of DD individuals have made the argument that variants inherited from unaffected parents can be diagnostic^6^. Additionally, recent studies of autism, schizophrenia, and bipolar disorder have indicated a role for rare, damaging variants in more common and complex neuropsychiatric disorders^11–17^. These rare variant burdens are most often concentrated in genes intolerant to loss-of-function variants in the general population (“constrained genes”) or those genes previously associated with severe DDs. Damaging variation in these two gene sets has also been tied to reduced educational attainment and cognitive performance in population cohorts such as the UK Biobank^18–23^, further strengthening evidence of their role in DDs.

Here, we sought to define the contribution of rare, and mostly inherited, variation to DDs in the DDD cohort. We jointly processed the entirety of the DDD cohort (n = 32,065) with relatively healthy UK population samples from the INTERVAL study (n = 3,943), which allowed us to evaluate both within-family and case/control rare variant burdens (**Fig 1a**). We found a significant burden of rare, damaging variants, particularly concentrated in constrained and known DD-associated genes, in both our case/control analysis and family-based analysis. In line with these results, we also found that the parents in the DDD cohort have a significantly higher burden of rare variants compared to controls. Finally, we combine gene-association evidence from our prior *de novo* study^24^ with the results of these analyses using TADA^13,25^, and nominate an additional 25 candidate DD genes for further investigation.

**Figure 1.**
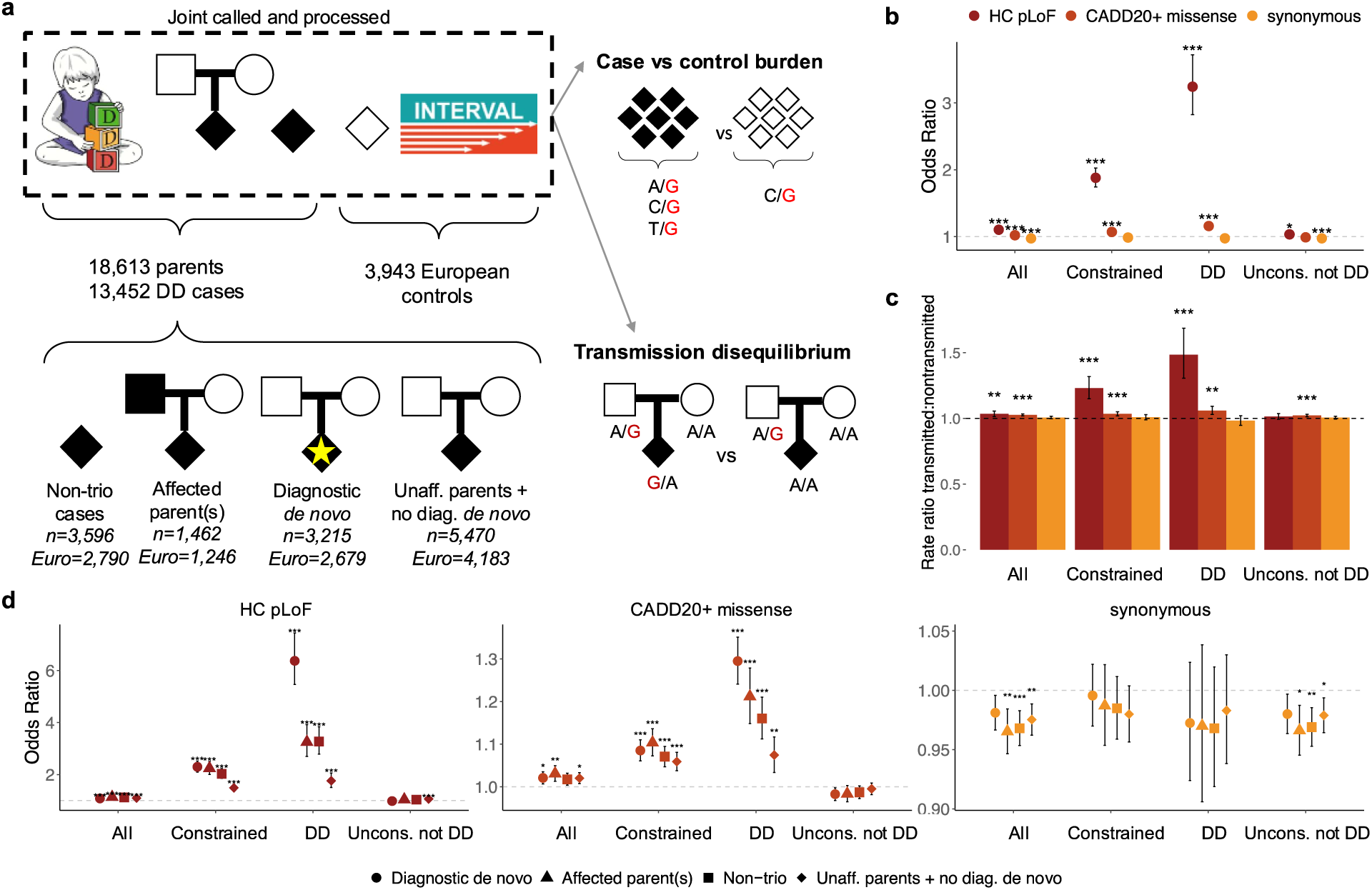
Overview of the dataset and analyses. a) Schematic representation of the study, showing the joint-calling of 32,065 individuals from the DDD study (18,613 parents and 13,432 DD cases) with 3,943 genetic-ancestry matched controls from the INTERVAL study. DD cases can be further subdivided into non-trio cases (n=3,596; 2,790 of inferred European genetic ancestry), trio cases with at least one affected parent (n=1,462; 1,246 of inferred European genetic ancestry), trio cases with a *de novo* diagnosis (n=3,215; 2,679 of inferred European genetic ancestry), and trio cases with unaffected parents and no *de novo* diagnosis (n=5,470; 4,183 of inferred European genetic ancestry). Given the joint-calling, we performed both case versus control burden testing and within-family transmission disequilibrium tests (TDT). Case/control burden (b) and transmission disequilibrium (c) testing for all DD cases (n=10,644 for case/control, 9,305 for TDT). d) Case/control regressions split by the type of DD case being compared to controls: trios with a diagnostic *de novo* variant are shown in circles, trios with one or more affected parents are shown with triangle, non-trio DD cases are shown in squares, and trios with unaffected parents and no *de novo* diagnosis are shown in diamonds. In b) and d), we report the odds ratios from logistic regressions. For the TDT tests in c), we are displaying the rate ratio between transmitted to nontransmitted variants. Displayed are the results for three mutation classes–LOFTEE high confidence predicted loss-of-function (HC pLoF, dark red), damaging missense with CADD ≥ 20 (orange), and synonymous (yellow) variants–and four gene sets: all genes (n=18,610), constrained (pLI ≥0.9, n=2,699), monoallelic DD-associated (n=666), and unconstrained genes with no prior monoallelic DD-association (n=15,667). * 1×10^-3^ ≤ p < 1×10^-2^; ** 1×10^-4^ ≤ p < 1×10^-3^; *** p < 1×10^-4^.

## Results

### Significant burden of rare variants in known DD-associated genes and constrained genes in DD cases

We joint-called autosomal SNVs and indels in the DDD cohort with controls from the INTERVAL study. After sample and variant quality control (see Methods), we retained 32,065 individuals from the DDD study (both parents and DD cases) of all inferred genetic ancestry groups. Of these individuals, 25,701 were of inferred European genetic ancestry for comparison against 3,943 ancestry-matched individuals from the INTERVAL study (**Supplementary Figure 1**). Post-quality control, we observed similar rates per exome of rare (allele frequency < 0.001) autosomal variants in DD cases, their parents, and controls (**Supplementary Table 1; Supplementary Figure 2**).

We first compared the rare (gnomAD^26^ allele frequency < 1×10^-5^; dataset allele frequency < 1×10^-4^) variant burden between DD cases and control individuals (n = 10,644 cases versus 3,943 controls), correcting for sex, the first 20 principal components (PCs), and the number of rare variants per exome, in line with recent work^12^. Correcting for the number of rare variants per exome leads to a significant odds ratio (OR) < 1 for synonymous variants when testing across all genes (OR = 0.974, p = 4.4×10^-6^, **Fig 1b**); we thus conclude that our ORs for damaging variants may be slightly under-estimated. Removing this correction, or correcting only for rare nonsynonymous variants, results in an OR significantly > 1 for synonymous variants that was not eliminated by various corrections (**Supplemental Note**).

We found modest ORs for the burden of rare predicted loss-of-function (pLoF) or damaging missense variants (defined as CADD^27^ ≥ 20) when evaluating all genes at once (LOFTEE^26^ high-confidence pLoF OR = 1.10 and p = 5.3×10^-19^, damaging missense OR = 1.02 and p = 5.3×10^-3^; **Supplementary Table 2**). However, these ORs are much larger in previously established monoallelic DD-associated genes^24,28^ (n=666 genes; pLoF OR = 3.24, p = 2.7×10^-63^) and constrained genes^26^ (pLI ≥ 0.9, n=2,699 genes; pLoF OR = 1.88, p = 1.0×10^-61^). We used all individuals in these analyses, but found nearly identical results when repeating the analyses using only unrelated individuals (**Supplementary Figure 3**). These results reinforce the role for these two gene sets in DDs, as has been demonstrated before^23,29^.

We then split DD cases into non-trio cases (n=2,790) and trio cases (n=7,854), which were further split based on whether (1) one or both parents were known to have a similar phenotypic presentation and (2) whether the proband had a likely diagnostic *de novo* variant. All groups have a significant burden of rare variants compared to controls (**Fig 1d**; **Supplementary Table 2**), with significantly higher ORs for pLoF variants in constrained and DD-associated genes compared to the set with unaffected parents and no *de novo* diagnosis (n=4,183; pLoF OR = 1.49 and 1.73, respectively) for the non-trio cases (pLoF OR = 2.03 and 3.27; Wald test p = 1.4×10^-6^ and 7.2×10^-8^, respectively), trios with a diagnostic *de novo* variant (n=2,679; pLoF OR = 2.30 and 6.37; Wald test p = 2.8×10^-11^ and < 1×10^-20^, respectively), and trios with one or more affected parents (n=1,246; pLoF OR = 2.42 and 3.25; Wald test p = 1.2×10^-8^ and 8.2×10^-7^, respectively; **Supplementary Table 3**). These significantly higher results were anticipated for the trios with one or more affected parents, where we expected that the affected parent would be transmitting risk variants to their children. For the trios with a likely diagnostic *de novo* variant, we might not have expected to see a large exome-wide burden given the presence of a high-impact variant (i.e., the diagnostic *de novo* variant), however this burden could be driven by the *de novo* variants.

Given that *de novo* variants play a large role in DDs and that they were included here, we repeated these regressions after removing the known *de novo* variants reported previously^24^ and observed an attenuated signal in all trio groups. This attenuation was most notable for the individuals that have a diagnostic *de novo* variant, which no longer have a significant burden of rare damaging variants compared to controls in all genes (pLoF OR = 1.00, p = 0.865; **Supplementary Figure 4**; **Supplementary Table 4**). However, this set of *de novo* diagnosed individuals does retain some signal in constrained (pLoF OR = 1.19, p = 3.9×10^-4^) and DD-associated genes (pLoF OR = 1.58, p = 8.2×10^-8^), even after removing the *de novo* variants.

While case/control comparisons are very sensitive to differences in genetic ancestry, within-family analyses are protected against such stratification, allowing us to test all trios. We sought to quantify the burden of rare variation that had been transmitted from parents using the Transmission Disequilibrium Test (TDT). We applied TDT to 9,305 DDD trios (all inferred genetic ancestry groups, but selecting only one sibling per family) and found a significant signal of overtransmission of rare (gnomAD allele frequency < 1×10^-5^; transmitted doubletons compared to nontransmitted singletons) damaging variants from parents to their children with DDs (**Fig 1c**; **Supplementary Table 5**). We found similar patterns as with the case/control analysis, with the strongest rate ratios (RRs) for pLoF variants in DD-associated and/or constrained genes (RR DD = 1.48 and p = 5.8×10^-10^; RR constrained = 1.23 and p = 1.4×10^-9^). Notably, we do not see any overtransmission of rare, synonymous variants in any of the gene sets tested (RR for all genes = 1.01, p = 0.183).

As in the case/control analysis, we split the trios by parental affected status and whether they had a diagnostic *de novo* variant. Even though the trios with one or more affected parents (n=1,306) had higher RRs than trios with unaffected parents and no *de novo* diagnosis (n=5,124) for pLoF variants in constrained (affected parents: RR = 1.51, p = 5.5×10^-7^; unaffected parents, no *de novo* diagnosis: RR = 1.28, p = 1.5×10^-7^; **Supplementary Table 5**) and/or DD-associated genes (affected parents: RR = 1.87, p = 1.7×10^-5^; unaffected parents, no *de novo* diagnosis: RR = 1.68, p = 2.1×10^-9^), the differences were not significantly different from each other (**Supplementary Table 6**). When evaluating the trios with a diagnostic *de novo* variant (n=3,155), we found no significant overtransmission of damaging variants in any of the gene sets (e.g., pLoF RR in constrained genes = 1.02, p = 0.800). These attenuated RRs for pLoF variants were nominally significantly lower than what was seen for unaffected parent and no *de novo* diagnosis trios, but not all comparisons survive multiple testing correction (p = 0.05/16 comparisons = 0.003; **Supplementary Table 6**). No differences were significant for missense variants between trios with a diagnostic *de novo* variant and the unaffected parent, no diagnostic *de novo* variant trios.

As both the TDT and case/control analyses establish, inherited rare variants significantly contribute to DDs as do common variants (see Huang*, Wigdor* et al.^9^). Given that we saw significant overtransmission of damaging variants from unaffected parents in the TDT results, we wanted to test if this was due to these parents having an excess of rare damaging variants compared to controls versus other potential explanations, such as ascertaining families where there was significant overtransmission of such variants. We therefore compared unaffected parents to controls with a similar logistic regression as above and found significant differences for missense and pLoF variation, albeit lower than what we found for the DD cases (13,861 unaffected parents versus 3,943 controls; **Supplementary Figures 5 & 6**). No significant difference was seen for synonymous variants. As expected, affected parents have higher burdens than unaffected parents, but the enrichments for pLoF variants in DD-associated and constrained genes are significant for both sets of parents (Fig 2a). While we found a few instances where mothers and fathers seemed to have a difference in their genetic burden – for example, the OR of pLoF variants in DD-associated genes in unaffected mothers was higher than in unaffected fathers (mother OR = 1.44 and p = 5.9×10^-4^, father OR = 1.05 and p = 0.643; Fig 2b) – these differences were not statistically significant (**Supplementary Figure 7**).

**Figure 2.**
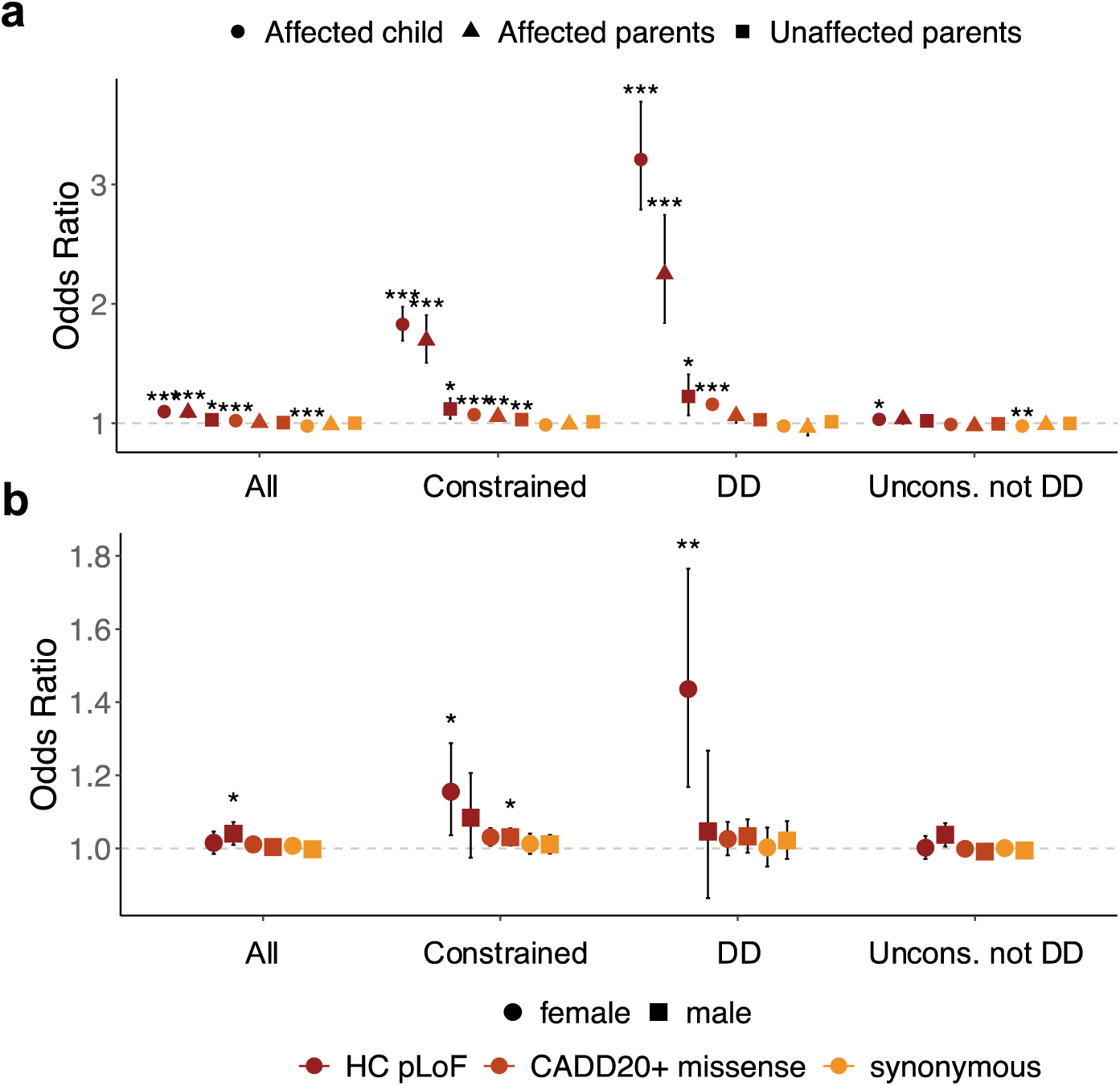
Parents of DD cases have significant rare variant burdens compared to controls. a) Odds ratio when comparing rare variant burdens to 3,943 controls for individuals of European genetic ancestry: developmental disorder cases in complete trios (“Affected child”, circles, n=7,854), affected parents (triangles, n=1,195), and unaffected parents (squares, n=13,861). b) The same plot as a), but with the unaffected parents split by sex of the parent. In all plots, high-confidence predicted loss-of-function (HC pLoF) variants are in dark red, damaging missense variants with CADD ≥ 20 are in orange, and synonymous variants are in yellow. There are four gene sets shown: all genes (n=18,610), constrained (pLI ≥0.9, n=2,699), monoallelic DD-associated (n=666), and unconstrained genes with no prior monoallelic DD-association (n=15,667). * 1×10^-3^ ≤ p < 1×10^-2^; ** 1×10^-4^ ≤ p < 1×10^-3^; *** p < 1×10^-4^.

### Investigating differences in rare variant burden

We wanted to further investigate differential burden in the DD cases, accounting for factors known to influence damaging genetic burden. Of note, neurodevelopmental disorders, particularly autism, have been reported to have a noticeable female protective effect^30,31^. The female protective effect would be expected to lead to patterns where female cases and mothers have a higher burden of rare, damaging variants compared to male cases and fathers, respectively. Indeed, we previously reported that female cases had a higher rate of damaging *de novo* variants compared to male cases^24,32^. However, we found no significant differences in the rare autosomal variant burden when comparing: (1) male to female cases (Fig 3a, **Supplementary Figure 8**), (2), unaffected mothers to unaffected fathers, as mentioned above (**Supplementary Figure 7**), or (3) the burden inherited from mothers to that inherited from fathers (**Supplementary Table 7**).

**Figure 3.**
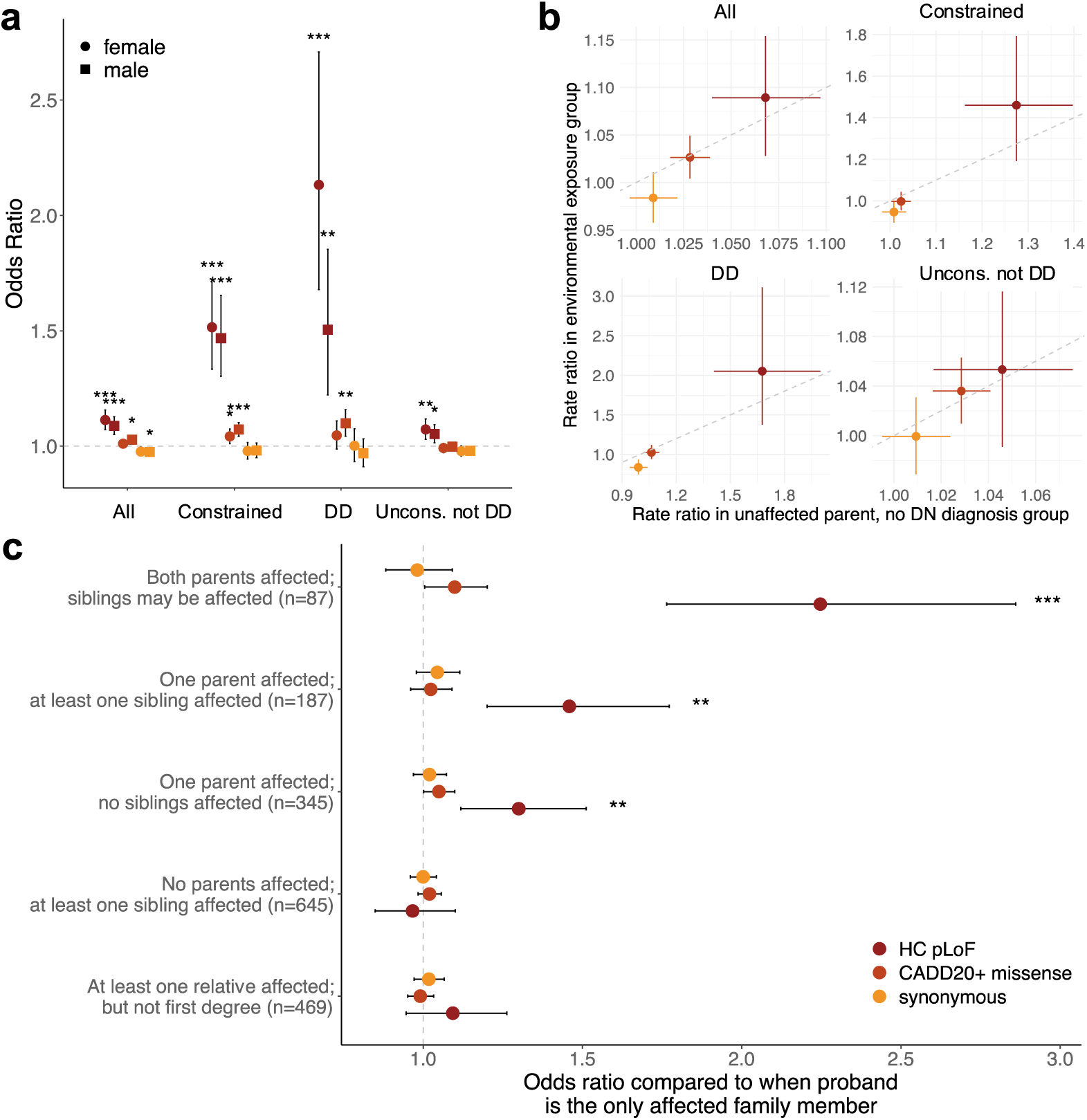
Comparison of rare variant burden between different subgroups of cases. a) Odds ratios when comparing female cases to female controls (circles, n = 1,651 and 1,940, respectively) and male cases to male controls (squares, n = 2,532 and 2,003, respectively) for individuals of European genetic ancestry with unaffected parents and no *de novo* diagnosis. While female cases have higher odds ratio estimates for high-confidence predicted loss-of- function (HC pLoF) variants in DD-associated genes, there are no significant differences for any group. b) The rate ratio from the transmission disequilibrium test (TDT) for cases of all genetic ancestries with unaffected parents and no *de novo* diagnosis (n = 5,124) compared to that set of cases who also had an environmental exposure (i.e., premature birth, maternal diabetes, and/or exposure to antiepileptic medications in utero; n = 1,075). There were no significant differences between the two sets. c) Odds ratios in constrained genes from a regression comparing unrelated European genetic ancestry cases with and without additional affected family members, split by the number and type of relative affected. All comparisons are against the burden in cases that are the only affected family member (n = 6,664). For all panels, LOFTEE high-confidence pLoFs are shown in dark red; missense variants with CADD ≥ 20 in orange; and synonymous variants in yellow. For a) and b), there are four gene sets shown: all genes (n=18,610), constrained (pLI ≥0.9, n=2,699), monoallelic DD-associated (n=666), and unconstrained genes with no prior monoallelic DD-association (n=15,667). Panel c) only shows constrained genes. * 1×10^-3^ ≤ p < 1×10^-2^; ** 1×10^-4^ ≤ p < 1×10^-3^; *** p < 1×10^-4^.

Under a liability threshold model, numerous genetic and environmental factors can contribute to the risk of developing a given disorder, and individuals whose accumulated risk factors (liability) cross a threshold are affected by the disorder^33^. The female protective effect discussed above can be thought of as one example of a liability threshold model, where, for example, females require a greater liability (e.g., burden of risk factors) than males to be diagnosed. Here, we wanted to explore other factors that could contribute overall to liability and would therefore influence the amount of expected burden from rare variation. For example, individuals who were born prematurely (< 37 weeks gestation) or had other environmental exposures in utero^34,35^ may be expected to have a greater liability coming from environmental factors and could have a lower rare variant burden than those without these exposures. In line with this, we have previously reported that this set of individuals has a lower diagnostic rate than those DD cases without prematurity of environmental exposures^2^. Similarly, DD cases who have affected family members have both been reported to have a significantly lower chance of having a *de novo* variant and of receiving a genetic diagnosis^2,36^. For those DD cases with affected family members, we would anticipate a greater inherited genetic burden of rare and/or common variants, the latter of which is explored in other work^9^, particularly given the lack of a large-impact diagnostic variant.

For environmental exposures, we selected a set of 2,637 probands within DDD who were born prematurely (< 37 weeks gestation), had mothers with diabetes, and/or were exposed to antiepileptic medications in utero, all three of which were shown to have lower diagnostic rates in DDD^2^. Rare variant burdens for DD cases with unaffected parents and no diagnostic *de novo* variant were similar whether they had or did not have an environmental exposure (Fig 3b, **Supplementary Figure 9**), with no significantly different burdens when directly comparing cases with (n=879) versus without (n=3,304) environmental exposures in a case versus case regression (**Supplementary Table 8**).

By contrast, when we compared rare variant burdens in unrelated European ancestry cases with versus without affected family members using a case versus case regression as above, we found a significant trend of increasing burden in constrained genes that was strongest for those cases with at least one affected parent (Fig 3c). These results are consistent with recent work^9^ that reported that cases with more affected first-degree relatives had lower polygenic scores for educational attainment^37^ as well as a lower chance of receiving a monogenic diagnosis. However, we did not see as strong of a trend for DD-associated genes (**Supplementary Figure 10**) despite this being the gene set that showed the largest difference from controls (**Supplementary Figure 6**), nor did we find any significant differences in rare variant burdens when the other affected family members were siblings or other first-degree relatives, even though these cases have a lower diagnostic rates and lower polygenic scores for educational attainment^9,36^.

### Burden of rare variation primarily resides outside of known DD-associated genes

Given the heterogeneity of DD cases within our cohort, we expected to be underpowered to detect significant burdens on an individual gene basis. When we performed a Fisher’s exact test to compare the burden of rare damaging variants in DD cases to those in controls, no individual gene crossed an exome-wide significance threshold (p < 2.8×10^-6^). The most significant gene was *ANKRD11* (pLoF carriers: 50 DD versus 1 control; Fisher’s OR = 18.6 and p = 3.7×10^-6^), a well-established DD-associated gene, with at least 20 *de novo* variants included in the DD carrier counts.

We therefore sought to estimate the excess burden in terms of the number of additional variants in DD cases in the significant gene lists from above. To do so, we estimated the excess of pLoF and damaging missense variants using the rate of observed variants in controls and corrected for the synonymous burden in each gene group. In the 4,183 DD cases with unaffected parents and no *de novo* diagnosis, we estimated an excess of approximately 4,000 pLoF and damaging missense variants, with two thirds coming from missense variants (**Supplementary Figure 11**). Only ∼11% of the excess was found in the previously DD-associated genes, indicating that there are more disease-associated genes to be identified. These excesses are not driven by specific individuals with far more variants than others, but by a minor shift in the distribution of the number of rare variants per individual (**Supplementary Figure 12**). Further, we estimated that the population attributable risk (PAR) from rare pLoF variants in constrained genes was 8.4%. This estimate of PAR was only minimally impacted by a range of population prevalence values for DDs, but drops to 5.9% when using the case/control analysis that removed known *de novo* variants (see **Supplemental Note** for caveats). This PAR estimate is similar to the 6% PAR reported for inherited pLoF variants in a recent autism study^15^.

To further investigate the nature of the rare variant exome-wide burden in the DD cases, we calculated *s_het_*burden scores^19^, a measure of the cumulative burden of rare variants made by combining the *s_het_* selection coefficients^38^ for each autosomal gene impacted by damaging variants (pLoF with CADD ≥ 25; missense with MPC^39^ ≥ 2 and CADD ≥ 25), for all individuals in the study (see Methods). Specifically, we wanted to evaluate if the exome-wide burden of rare inherited damaging variants in DD cases differed from the burden seen in parents. For these analyses, we removed known *de novo* variants from the DD cases when calculating *s_het_*burden scores. We found a significant difference in *s_het_* burden scores calculated from rare pLoF variants (CADD ≥ 25 as in Gardner et al.^19^) when comparing DD cases to the parental data (Wilcox p = 2.01 x 10^-9^), but not for *s_het_* burden scores calculated using synonymous variants (Wilcox p = 0.121; **Table 1**). This suggests an overtransmission of pLoF *s_het_* burden scores from parents to their children with DDs. Notably, we did not find a significant difference for *s_het_* burden scores calculated using damaging missense variants as defined by Gardner et al.^19^, likely as the class is too rare for there to be large differences (CADD ≥ 25 and MPC ≥ 2; **Supplementary Table 9**). We also found no significant differences between *s_het_* burden scores for male versus female DD cases, in line with analyses reported above.

**Table 1.** Differences in the *s_het_* burden scores for DD cases compared to their parents.

| Group | N | Synonymous |  | pLoF |  |
| --- | --- | --- | --- | --- | --- |
|  |  | Effect size<br>(95% CI) | P-value | Effect size<br>(95% CI) | P-value |
| All | 9305 | 0.0038<br>(-0.0042 – 0.0106) | 0.121 | 0.0045<br>(0.0027 – 0.0064) | 1.88e-09 |
| European | 7037 | -0.0029<br>(-0.0108 – 0.0064) | 0.732 | 0.0047<br>(0.0028 – 0.0067) | 6.01e-08 |
| Unaffected parents,<br>no DNM diagnosis | 5124 | 0.0056<br>(-0.0048 – 0.0151) | 0.344 | 0.0047<br>(0.0023 – 0.0074) | 2.37e-05 |
| DNM diagnosis | 3155 | 0.0052<br>(-0.0093 – 0.0214) | 0.111 | 0.0022<br>(-0.0004 – 0.0049) | 0.002 |
| Affected parents | 1306 | -0.0093<br>(-0.0262 – 0.0092) | 0.992 | 0.0091<br>(0.0035 – 0.0152) | 0.001 |

Shown are *s_het_* burden scores made when using rare synonymous variants or rare pLoF variants with CADD ≥ 25 for five sets of trios. In the case where a family had multiple affected children, only one child (and thereby complete trio) was used. For these analyses, all known *de novo* variants were removed from calculations. Bootstrapping with 1000 replicates was used to determine the median difference in *s_het_* burden scores as well as the 95% confidence intervals (CI). P-values are from the Wilcoxon rank sum test.

We additionally compared the *s_het_* burden scores between unrelated European genetic ancestry DD cases (n=8,062) and controls in a logistic regression, which allowed us to estimate the variance explained on the liability scale. Assuming a population prevalence of 1% for DDs, we estimated that the *s_het_*burden scores explained 2.6% [0.2 - 7.5%] of the variance on the liability scale, which is lower than the estimated 11.2% due to common variants genome-wide^9^. Additionally, we found that the variance explained was higher if only using the *de novo* diagnosed individuals (n=1,905 cases; 4.6% of liability [1.1 - 10.1%]) compared to only undiagnosed DD cases (n=6,157; 1.9% [0.4 - 6.3%]). By contrast, the raw counts of rare pLoF and damaging missense variants would only explain 0.06% [0.03 - 0.1%] of the variance on the liability scale.

## Combining evidence from *de novo* and inherited variants identifies additional candidate DD genes

We previously identified nearly 300 genes significantly associated with DDs via a gene-specific enrichment of *de novo* variants^24^, with dozens more genes near the significance threshold (“31k analysis”). We wanted to combine evidence from *de novo* variants with the TDT and case/control analyses performed here to improve our ability to identify DD-associated genes. We applied the Transmission and De Novo Association (TADA) framework^13,25^, a Bayesian analytical framework that makes use of priors on the risk of a given variant class in each gene to measure statistical evidence, which has been successfully applied to studies of the genetic basis of autism^13,25,40^.

At an exome-wide significance threshold of 2.8×10^-6^, we identified 269 significant genes (**Supplementary Table 10**). As in the prior autism studies, pLoF variants had the largest combined Bayes Factor (BF) contribution for the variant types tested (66.7%; Fig 4a), and we found that nearly all of the BF contribution came from *de novo* variants (97.2%; Fig 4b) with case/control a distant second (2.5%). Unsurprisingly given the strong *de novo* contribution, the vast majority of these 269 genes were significant in our previous 31k analysis^24^ (217; 81%). To identify a set of genes with limited prior association to DDs, we focused on the 52 genes that were not significant in the prior study of *de novo* variants. Of those, 27 already had strong or definitive evidence of being monoallelic DD-associated genes according to DDG2P^28^ as of July 2023 (**Supplementary Figure 13**). We selected the remaining 25 genes (2 with moderate, 8 with limited, and 15 with no evidence of being monoallelic DD-associated genes in DDG2P) for deeper investigation (Fig 4c). For most of these genes, the primary driver of association was the signal from *de novo* pLoF variants, although we saw stronger case/control and inherited signals as measured by BF contribution for these 25 genes compared to the 244 significant but known genes (Fig 4d**-e**; **Supplementary Table 11**).

**Figure 4.**
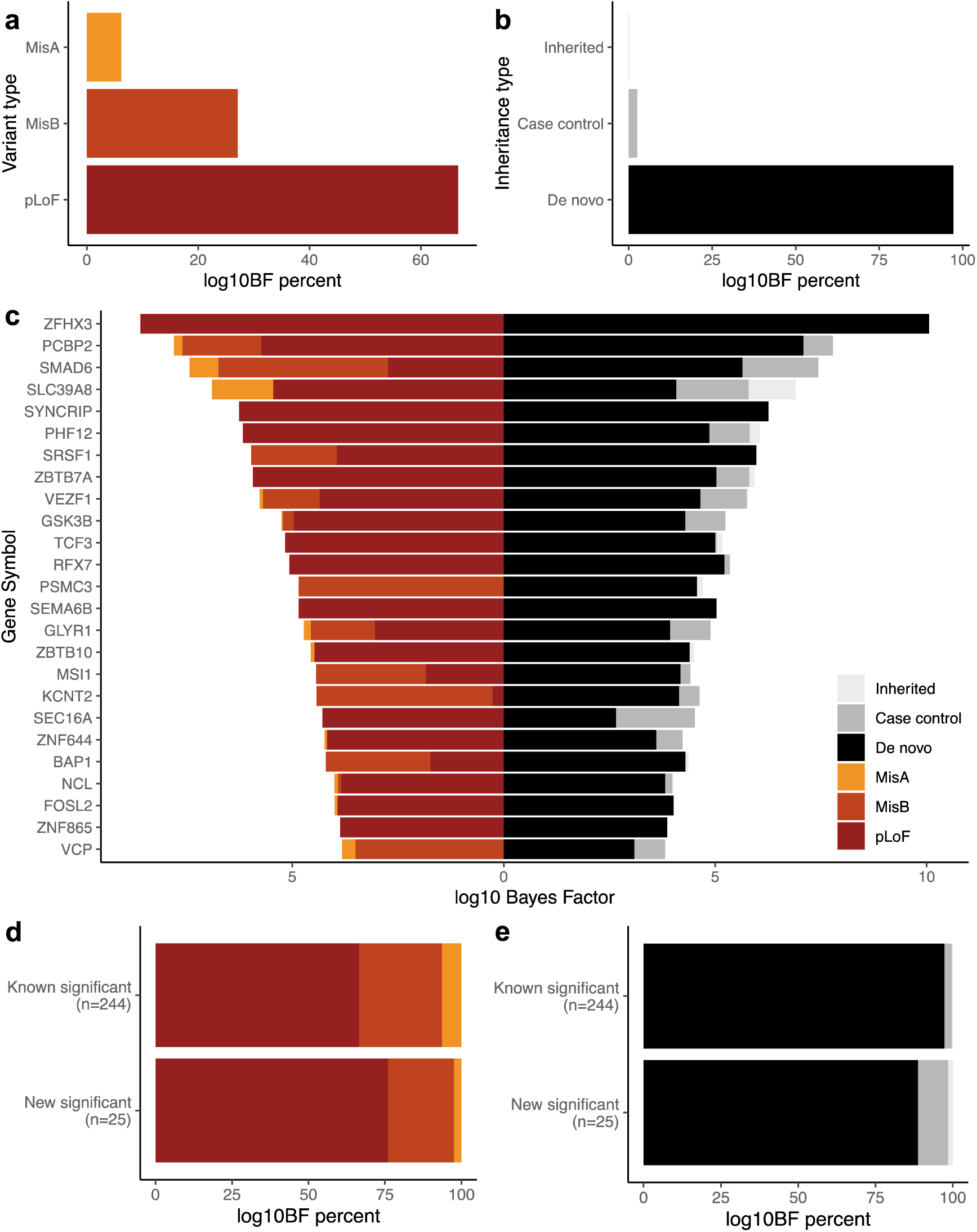
Results from a combined *de novo*, case/control, and inherited analysis of rare variants in developmental disorder (DD) cases using the Transmission and De Novo Association (TADA) framework. The evidence of association with DD, as measured by Bayes Factors (BF) in TADA, for different variant types (a) and inheritance types (b) across all 269 significant genes (p < 2.8×10^-6^). In c) we depict the specific Bayes Factors from TADA for 25 genes that were significant in the TADA analysis, but were not significant in our prior analysis of *de novo* variants alone^24^ and had limited or no evidence of DD association in DDG2P. In d) and e) the Bayes Factor percentages are compared for the TADA significant genes with previous DD-associations (“Known significant”, n=244 genes) versus those with minimal prior evidence DD-association (“New significant”, n=25 genes) for different variant types (d) and inheritance types (e). For all plots, high-confidence predicted loss-of-function (pLoF) variants are depicted in red, missense variants with MPC ≥ 2 (misB) in orange, and missense variants with 1≤ MPC < 2 (misA) in yellow. Similarly, signal from *de novo* variants are depicted in black, case/control in dark gray, and inherited (e.g., transmitted versus nontransmitted) in light gray.

For these 25 genes with limited evidence of association in DDG2P, we further classified them as having high, medium, or low likelihood of being true DD-associated genes based on manual curation of additional gene-phenotype databases (e.g., OMIM^41^, GenCC^42^, PanelApp^43^), constraint scores^26,44,45^, literature searches, and evaluation of the Bayes Factors from TADA (see **Supplemental Note** for more details about classification; **Supplementary Figures 14-15**). Eleven of these 25 genes had strong evidence of being *bona fide* DD-associated genes, primarily due to being considered by other clinical centers to have strong enough evidence to use for diagnosis in closely related phenotypes (e.g., *SYNCRIP*) or recent publications (e.g., *FOSL2*^46^). Thirteen of the genes were considered to have medium evidence of DD association; the majority of these genes had strong statistical evidence but no additional evidence in the literature. We note that 77% of these genes are highly constrained against loss-of-function variation in the general population (10 of 13 genes are LOEUF < 0.35), a feature that is greatly upweighted in TADA. Finally, one gene, *GLYR1*, had weaker evidence for true DD-association given that two of the three *de novo* pLoF variants, which were the primary driver of association, were found in patients that could be considered diagnosed by other variants. These findings diminish the strength of association, but as a small subset of DDD cases have dual molecular diagnosis^2^, we cannot fully rule out this gene as a potential DD-associated gene.

For the eleven genes with a high likelihood of being true DD genes (*BAP1*, *FOSL2*, *KCNT2*, *PSMC3*, *RFX7*, *SEMA6B*, *SRSF1*, *SYNCRIP*, *VCP*, *ZBTB7A*, *ZFHX3*), we investigated how many of individuals harboring damaging variants in these genes were considered diagnosed with other variants (see **Supplemental Note** for details). When using a Bayes Factor threshold of >1.3 to define variant carriers, we found that only 14.1% (12 of 85) of individuals with one of these variants has some other diagnosis, which was significantly lower than the overall diagnostic rate (27.5%; Fisher’s exact p = 4.8×10^-3^). Of note, we recently estimated^47^ that a similar percentage (∼12.5%) of DDD cases diagnosed with a *de novo* variant could have a second *de novo* diagnosis, with most of that burden outside of the known genes. These results emphasize that these eleven genes are more likely to be *bona fide* DD-associated genes.

Surprisingly, we found that a known biallelic DD-associated gene, *SLC39A8*, was significant in the TADA analysis (p = 1.14 x 10^-9^) with contributions from all three inheritance classes (i.e., *de novo*, case/control, and inherited). We reported a similar set of findings for *KDM5B* previously^48^, which also had signal from *de novo*, biallelic, and overtransmission of pLoFs. *SLC39A8* shows some depletion of both pLoF and missense variants in population cohorts (gnomAD^26^ v2: pLoF observed / expected = 0.18 and missense observed / expected = 0.8), but does not have particularly strong constraint scores (pLI = 0.67 and LOEUF = 0.47), meaning that the priors in TADA would not be large. Focusing on the contribution from pLoF variants to the association signal for *SLC39A8*, there were three individuals with a *de novo* pLoF in *SLC39A8*, four non-trio individuals that contributed to the case/control analysis (with zero found in controls), and eight instances where a parent transmitted a pLoF variant (as part of an 8:0 transmitted:nontransmitted signal). Across these fifteen individuals, only one carried another putatively diagnostic variant – one of the individuals with a *de novo* pLoF variant in *SLC39A8* also harbored a *de novo* missense variant in *DDX6*, a known DD-associated gene. For those individuals in the DDD cohort (all twelve in the case/control and inherited analysis), we searched for additional rare coding variants in *SLC39A8* that could contribute to a recessive diagnosis. As both missense and pLoF variants have been reported as pathogenic or likely pathogenic in ClinVar^49^ for *SLC39A8*, we included both in our search, but found no additional rare variants in the twelve patients we queried.

Additionally, we noted a striking 24:3 transmitted:nontransmitted signal for missense variants with moderate MPC scores^39^ (1 ≤ MPC < 2) in *SLC39A8*. Three of these 24 individuals had a partial or full diagnosis from other variants, but the rest were considered undiagnosed (n=21; 87.5%). As above, we searched for other rare coding variants in *SLC39A8* and found a second missense variant inherited from the other parent for only one individual, whose phenotypes were consistent with the recessive SLC39A8-Congenital Disorder of Glycosylation^50^. The remaining individuals have no other convincing diagnostic candidate or other rare coding variants in *SLC39A8*. Based on these analyses, we consider *SLC39A8* a promising candidate DD-associated gene that can operate via both monoallelic and biallelic mechanisms, like the previously reported *KDM5B*.

## Discussion

In this work, we demonstrated a significant contribution of rare, typically inherited, damaging variants to the risk of severe developmental disorders (DD) by comparing both the rare variant burden in DD cases to controls, as well as evaluating within-family overtransmission of these variants from parents to their affected children. The majority of this rare variant burden was found in genes intolerant of loss-of-function variants in reference population datasets (i.e., constrained genes)^26^ and genes previously associated with DDs via monogenic forms of inheritance, as has also been seen for other neurodevelopmental and psychiatric disorders^11–13,17,29^. We estimate that rare pLoF variants in constrained genes, for example, account for 6- 8.5% of the population attributable risk in our cohort. While the burden of overtransmission was stronger from affected parents, cohort-wide nearly all of the burden was transmitted from unaffected parents (e.g., 92.5% of transmitted rare pLoFs and missense variants across all genes are from unaffected parents). In a set of DD cases with unaffected parents, we found an excess of thousands of rare pLoF and damaging missense variants spread across many genes and concentrated outside the previously DD-associated genes, indicating that there are additional DD risk loci to be identified. Finally, we applied the Transmission and De Novo Association (TADA) framework^13^ to combine the evidence of association from *de novo* variants, case/control analyses, and the overtransmission of variants within families, and identified 269 significant genes, of which 25 had limited to no prior evidence of DD-association.

When removing *de novo* variants, which are known to be large contributors to DD risk^24^, from our case/control analyses, we still found significant burdens of rare pLoF and, to a lesser extent, damaging missense variants in constrained and DD-associated genes across all DD cases (**Supplementary Figure 4**), even in those with a likely diagnostic *de novo* variant. In recent work, we report that DD cases with a monogenic diagnosis have higher common, polygenic risk than controls, but significantly less risk than undiagnosed cases^9^. However, this increased polygenic risk was driven nearly entirely by affected parents in the diagnosed cases. When repeating the case/control analyses for trios with a diagnostic *de novo* variant and removing those with an affected parent (n=2,425 DD cases remaining), we found significant burdens of rare pLoF and missense variants (**Supplementary Table 12**). Similarly, recent work in DDD has shown via burden analysis that ∼12.5% of probands who currently have a single molecular diagnosis may also have an additional *de novo* variant (mostly outside of known genes) contributing to their condition^47^. These results show that rare, inherited variants may be contributing both to DD risk and to phenotypic presentations for DD cases both with and without a monogenic diagnosis.

The significant overtransmission of rare pLoF and damaging missense variants from parents to their children with DDs could arise via multiple mechanisms, including skewed transmission of deleterious variants and/or an overall excess of such variants in the parents themselves. When directly testing the latter, we found that parents of DD cases–even those apparently unaffected by DD-related phenotypes–were significantly different from ancestry-matched controls in terms of their rare autosomal variant burden (Fig 2). The ORs were attenuated compared to the DD cases, but were still significant for pLoF variants in constrained and/or DD-associated genes (**Supplementary Figure 5**). While we did not see a significant difference between unaffected mothers and unaffected fathers (**Supplementary Figure 7**), as we might have predicted based on previous work on sex differences in reproductive success^19^, we had incomplete power to detect modest differences. Beyond suggestive differences in the ORs (e.g., in Fig 3a comparing male and female cases), we could not find any significant sex-specific differences in rare variant burdens in this cohort, in line with the recent findings from a smaller cohort of intellectual disability^29^. Similarly, while we have reported a higher burden of *de novo* variants in female cases previously^24,51^, this excess was only found in well established DD-associated genes with no significant differences in the *de novo* burden between male and female cases in genes not associated with DDs or in undiagnosed cases^24^.

Beyond diagnostic status and sex, we investigated other factors that could be associated with differential rare variant burden within our cohort. While we found no differential burden for DD cases with an environmental exposure, we found a relationship between genetic burden and the number of family members with similar clinical phenotypes as in recent work on polygenic burden^9^. Here, we observed stronger rare variant burdens in constrained genes (Fig 3c) as the number of affected relatives increases, which is consistent with recent findings from Huang*, Wigdor* et al. when comparing polygenic scores for traits related to DD-risk, such as educational attainment^9,37^. However, we did not see this same trend for DD-associated genes (**Supplementary Figure 10**), in contrast to their work and that from Urpa and colleagues^29^.

We estimated that there was an excess of ∼4,000 rare pLoF and damaging missense variants in 4,183 DD cases with unaffected parents and no diagnostic *de novo* variant, and that only ∼10% of this excess could be found within the known monoallelic DD-associated genes (**Supplementary Figure 11**), which prompted us to search for additional DD risk genes. Given our lack of significance for individual genes in a per-gene burden test and the knowledge that *de novo* variants play a substantial role in DDs, we applied the TADA Bayesian framework to combine association evidence across multiple variant classes (e.g., *de novo* and inherited), following work from studies of autism^13^. Nearly all of the 269 significant genes from TADA were previously tied to DDs either via our prior study on *de novo* variants^24^ or the manually-curated DDG2P^28^ list. Of these significant genes, 25 had minimal evidence for prior association as of July 2023 and eleven have additional evidence supporting their association with DD based on recent publications or their addition to other DD-related gene lists. These 25 genes, however, only represent ∼1.5% of the excess of rare pLoF and missense variants reported above, again reinforcing that there are additional DD-associated risk loci to be discovered.

In the TADA analysis, we found that the major driver of association was the *de novo* variant contribution (Fig 4b). Indeed, ∼80% of the significant genes in the TADA analysis were significant in our prior study of *de novo* variants alone^24^ and, of those that were not significant, nearly all had suggestive evidence of association in the prior study (e.g., 20/25 with p-values < 0.001). For these genes with suggestive evidence previously, the inclusion of case/control and inherited data provided enough additional statistical evidence to cross an exome-wide significance threshold. In fact, the 25 TADA significant genes with minimal prior DD-association evidence have a larger contribution from case/control or inherited analyses in the TADA framework (Fig 4d**-e**; **Supplementary Table 11**) compared to the previously DD-associated genes. For the eleven genes that were determined to have a high likelihood of being *bona fide* DD-associated genes, we found that carriers of damaging variants in these genes were clinically diagnosed at a significantly lower rate than the cohort overall (14% versus 27.5%, p = 4.8×10^-3^). In some of the genes with mixed evidence of being *bona fide* DD risk genes (e.g., *ZNF644*), the association signal seemed to be driven primarily by the genes’ low LOEUF scores (indicating severe selective constraint; **Supplementary Figure 15**), which are strongly upweighted in the TADA framework (**Supplementary Table 13**). A deeper analysis of the phenotypic profiles of individuals with damaging genetic variation in these genes would provide more confidence in their association with DDs.

Beyond the limitations of sample size–which is most notable when the cohort is subdivided by diagnostic status and parental affected status–another limitation of this study is the inability to know which variants are truly having a large functional impact. We aimed to reduce this limitation by using strict allele frequency thresholds and filtering only to LOFTEE high-confidence pLoF variants and missense variants with some evidence of being deleterious (e.g., CADD ≥ 20). However, we know that both rarity and *in silico* score deleteriousness predictions are not sufficient to establish true variant impacts. For example, manual curation of variants from the gnomAD database has shown that a large fraction of pLoF variants in haploinsufficient genes associated with severe phenotypes have evidence that they are not true loss-of-function variants^52,53^. Generally, there is a need to better understand the incomplete penetrance of DD-risk variants, and analyses like those presented here would be improved by the creation of approaches to estimate penetrance of risk variants from population cohorts with longitudinal health data. Finally, we did not consider combinations of rare variants or the interaction between rare and common variants, but a recent paper by Urpa et al. found evidence of additivity between rare and common variants when studying individuals with intellectual disability^29^. A more integrative model that could account for the contributions of rare variants, polygenic risk, and environmental factors would further improve assessment of DD risk.

While this study has established a role for rare, inherited variants in DD risk, it will do little to improve the diagnostic rate of patients with DD primarily because these risk-increasing variants are often inherited from apparently unaffected parents. The majority of diagnostic pipelines do not consider variants inherited from unaffected parents, even with increasing evidence that incompletely penetrant variants can contribute both to diagnoses^6,10^ and to related phenotypes^23^. Diagnostic pipelines could be modified to allow for the consideration of these lower penetrance variants, but such a change would need to balance the inclusion of these potential risk variants with additional clinical curation time to evaluate the many other variants that would also qualify for diagnostic consideration (e.g., ∼19% of the trios have inherited at least one rare pLoF variant in a constrained gene from an unaffected parent).

Potentially deeper phenotyping of the parents–including collecting data on their educational history and phenotypes throughout life–would reveal sub-clinical phenotypes or phenotypes that were stronger earlier in life that are not readily apparent in routine interactions with clinicians when enrolling their children in studies such as DDD. Indeed, concurrent work in birth cohorts (Malawsky et al. *in prep*) has shown that rare damaging variants in constrained genes have a larger impact on cognitive ability early than later in life, implying that the parents may have had learning difficulties in early childhood that subsequently improved. Considering sub-clinical or earlier life phenotypes of parents could aid in identifying or prioritizing inherited risk variation in DD cases, but would also necessitate additional genetic counseling considerations, such as the impact on recurrence risk and the potential disclosure of these incompletely penetrant variants to the parents.

There is also the possibility that parents appear to be able to tolerate apparently damaging genetic variation due to other genetic, environmental, or stochastic factors that either protect the parents or increase susceptibility in their children. For genetic factors, it has been suggested that protective polygenic backgrounds (e.g., higher overall educational attainment polygenic scores) or *cis*-regulatory variation that reduces the penetrance of the damaging genetic variant^54^ could contribute. While Kingdom et al.^23^ reported evidence of educational attainment polygenic scores protecting against the phenotypic impacts of carrying rare, damaging genetic variants, we found no evidence of this in a study of 11,573 DD cases^9^. We also found limited evidence for the protective effect of *cis*-regulatory variation in a set of 1700 trios where DD cases inherited rare, putatively damaging variants from unaffected parents^55^.

Taken together, our findings indicate that most, if not all, DD cases in our cohort have genetic contributions to risk from rare inherited variation, even those individuals with an established diagnosis from a presumably large-effect *de novo* variant. We found stronger rare variant burdens in undiagnosed cases and those with affected family members, as anticipated given concurrent work on the contribution of polygenic scores^9^ to DDs and prior study of the factors that impact diagnostic rates^2^. However, there is still much work to be done to both identify additional DD-associated genes and to understand how these rare variants are increasing risk for DD, including how they may interact with polygenic risk and environmental risk factors. Larger sample sizes, particularly those with access to longitudinal and comprehensive phenotype or health record information for the parents, will improve our understanding of the genetic architecture of DDs and will provide more insight into the mechanisms of incomplete penetrance for rare, inherited damaging genetic variation.

## Data Availability

Sequence and variant-level data and phenotype data from the DDD study data are available on the European Genome-phenome Archive (EGA; https://www.ebi.ac.uk/ega/) with study ID EGAS00001000775. Exome sequencing for the INTERVAL cohort is also available on EGA with study ID EGAD00001002221. Previously described databases were from the Genome Aggregation Database (gnomAD v2.1.1; https://gnomad.broadinstitute.org/downloads) and the Developmental Disorders Genotype-Phenotype Database (DDG2P; https://www.ebi.ac.uk/gene2phenotype/downloads).

## Acknowledgements

We thank the families and their clinicians for their participation and engagement, and our colleagues who assisted in the generation and processing of data. The DDD study presents independent research commissioned by the Health Innovation Challenge Fund (grant number HICF-1009-003). The full acknowledgements can be found at www.ddduk.org/access.html. We additionally thank Rachel Hobson and Rosemary Kelsell for DDD project management, and the Human Genetics Informatics (HGI) group at Sanger and Aleksejs Sazonovs for fruitful discussions and support during quality control of the data. This study makes use of DECIPHER, which is funded by the Wellcome Trust. This research was funded in part by Wellcome (grant no. 220540/Z/20/A, “Wellcome Sanger Institute Quinquennial Review 2021–2026”). For the purpose of open access, the authors have applied a CC-BY public copyright license to any author accepted manuscript version arising from this submission.

## Author Contributions

K.E.S. performed analyses and figure generation. K.E.S., V.K.C., E.J.G., P.D., E.M.W., S.J.L., T.S., R.Y.E., and G.G. were involved with data generation and quality control. V.K.C., J.M.F., D.S.M, S.J.L., and P.C. contributed to code, methods, or additional data. The DDD study is supervised by C.F.W., H.C.M., H.V.F., and M.E.H. M.E.H. supervised this study. The primary writing was completed by K.E.S. with input from V.K.C., C.F.W., H.C.M., and M.E.H. All authors approved the final manuscript.

## Code Availability

Analyses were primarily performed with Python and R (version 4.2.0). The R code used to generate the TADA results are available at: https://github.com/talkowski-lab/TADA_2022

## Methods

### Samples included in analyses

#### Deciphering Developmental Disorders (DDD)

Patients with severe, undiagnosed developmental disorders were recruited from 24 regional genetics services within the United Kingdom National Health Service and the Republic of Ireland between 2011 and 2015. Families gave informed consent to participate, and the study was approved by the UK Research Ethics Committee (10/H0305/83 granted by the Cambridge South Research Ethics Committee, and GEN/284/12 granted by the Republic of Ireland Research Ethics Committee). The inclusion criteria included neurodevelopmental conditions, congenital, growth or behavioral abnormalities, and dysmorphic features. Additional details on sample collection, exome sequencing, alignment, variant calling (inherited and *de novo*) and variant annotation have been described previously^24,51^. In brief, exome capture was carried out with either Agilent SureSelect Human All Exon V3 or V5 baits. Reads were aligned to the GRCh37 1000 Genomes Project phase 2 reference (hs37d5) using BWA aln and BWA mem^56,57^. Variants were called using GATK’s HaplotypeCaller, CombineGVCFs, and GenotypeGVCFs (GATK version 3.5.0)^58^, and then restricted to merged bait regions from the two capture kits plus 100 base pairs of padding on either side.

After removing samples that had withdrawn consent for research, these analyses involve 13,452 individuals with developmental disorders: 9,856 individuals with complete trios from 9,305 families, and 3,596 non-trio individuals. *De novo* and inherited variants from a subset of these individuals have been published previously^24,48^.

#### Controls from the INTERVAL study

The INTERVAL study^59^ was a randomized controlled trial of the safety and efficacy of varying the duration between blood donations^60^. As part of this work, 50,000 presumed healthy adults (18 years or older) were consented and recruited from NHS Blood and Transplant blood donation centers across England. DNA extraction and genotyping was funded by the National Institute of Health Research (NIHR), the NIHR BioResource (http://bioresource.nihr.ac.uk/) and the NIHR Cambridge Biomedical Research Centre (www.cambridge-brc.org.uk). The academic coordinating center for INTERVAL was supported by core funding from: NIHR Blood and Transplant Research Unit in Donor Health and Genomics, UK Medical Research Council (G0800270), British Heart Foundation (SP/09/002) and NIHR Research Cambridge Biomedical Research Centre. A complete list of the investigators and contributors to the INTERVAL trial is provided in Moore et al.^59^ and at www.intervalstudy.org.uk/about-the-study/whos-involved/interval-contributors/.

For a subset of the INTERVAL cohort, exome sequencing was performed as described previously^61,62^ using the Agilent SureSelect Human All Exon V5 baits. In this study, we had access to exome sequencing data from 4,502 individuals. As detailed below, 3,943 of these individuals were retained for analyses after QC and selecting for inferred northwestern European genetic ancestry.

### Data quality control

#### Variant quality control

We performed quality control on autosomal single nucleotide variants (SNVs) and insertions/deletions (indels). We tested a range of values for both genotype-level (e.g., genotype quality) and variant-level metrics (e.g., VQSLOD).

For SNVs, we selected our quality control filters by evaluating (1) the transmission rate of rare synonymous variants from a parent to their child (allele count [AC] = 2, only seen in parent and their child) and (2) sensitivity to retaining known *de novo* variants^24^. The final filters for SNVs were:

- VQSLOD ≥ -2
- Genotype quality (GQ) ≥ 20
- Depth (DP) ≥ 7
- p-value for sampling the observed allele balance under a binomial model, assuming an allele balance of 0.5 for heterozygous sites > 1×10^-3^
- Fraction of non-missing genotypes passing genotype-level QC thresholds > 0.5

Using the above thresholds, our transmission rate of singleton synonymous variants was 0.500 (e.g., perfect balance of transmitted to nontransmitted variants) and our sensitivity to recovering known *de novo* variants was 88%. Analyses were performed with bcftools^63^.

For indels, we selected our quality control filters by evaluating (1) the transmission rate of rare inframe variants in unconstrained (pLI < 0.9), non-monoallelic DD-associated genes from a parent to their child, (2) the sensitivity to retaining known *de novo* variants, and (3) the frameshift:nonsense ratio, which we expected to be 1-1.2. The final filters for indels were:

- VQSLOD ≥ -2
- GQ ≥ 25
- DP ≥ 10
- Allele balance (AB) > 0.3

Using the above thresholds, our transmission rate of rare inframe variants in unconstrained, non-monoallelic DD-associated genes was 0.490 (not significantly different from the expected 0.5), our sensitivity to recover known *de novo* indels was 72%, and our frameshift:nonsense ratio was 1.10. Finally, we removed indels found in the same gene and sample, which represented ∼4% of all indels with minor allele frequency (MAF) < 1%. These indels were often part of a complex mutational event that would require haplotype-aware annotations to resolve.

#### Variants annotation and filtering

Variants were annotated with VEP^64^, including the LOFTEE plugin^26^. We additionally annotated all variants with CADD v1.4^27^ and MPC^39^.

Finally, variants in the joint VCF were filtered using the following criteria:

- gnomAD v2.1 raw allele frequency < 0.001
- Coding consequences based on VEP annotations in protein-coding genes
- On autosomes

Note that nearly all analyses in this work use a lower gnomAD allele frequency, namely MAF < 1×10^-5^, as well as a filter on dataset allele frequency. Additionally, we used the worst consequence in protein-coding transcripts as the mutational consequence for variants.

#### Sample relatedness

KING^65^ was run to determine relatedness between samples in this joint-called cohort. We used common variants (MAF > 1%) that passed the SNV filters listed above and, after applying those filters, had low missingness (< 5%). Related individuals were defined as those with a kinship coefficient > 0.04419417, which is the lower bound cut-off for third-degree relatives. A list of unrelated parents (n = 18,494) and probands (n = 10,613) was created to maximize the number of samples retained.

### Determining a set of individuals with inferred European genetic ancestry

For analyses comparing cases to controls, we needed to identify a subset of individuals who had similar inferred genetic ancestries. Given that both DDD and INTERVAL were primarily collected in the United Kingdom, our largest genetic ancestry group for comparison was individuals with inferred northwestern European genetic ancestry (e.g., matching ancestry historically tied to the British Isles).

After splitting multiallelic variants, we selected common (MAF > 1%) single nucleotide polymorphisms (SNPs) that passed QC filters (GQ ≥ 20 and DP ≥ 7) and had < 10% genotype missingness. Ambiguous SNPs and indels were removed as were variants in 24 long-range linkage disequilibrium (LD) regions, such as the HLA^66^. Overlapping SNPs from the 1000 Genomes Phase 3 individuals^67^ were identified and filtered to those with MAF > 1% and genotype missingness > 10%. SNPs were then merged between our DDD/INTERVAL joint-call and the 1000 Genomes, and LD-pruned (r^2^ < 0.2), leaving 32,413 variants. Principal component analysis (PCA) was performed on the 1000 Genomes samples using the smartpca function from EIGENSOFT^68,69^, with the DDD and INTERVAL samples projected onto the resulting PCs.

Further dimensionality reduction was done using UMAP on the first 20 PCs, and the individuals of European genetic ancestry were defined as those overlapping with the labeled European ancestry individuals from the 1000 Genomes Project.

We needed to generate PCs within the inferred European genetic ancestry subgroup to use as covariate in the case/control regressions. To do this, we extracted variants from a set of unrelated parents and controls as defined by KING^65^ with inferred European genetic ancestry. We performed similar variant filtering as above (e.g., MAF > 1%, passing QC, not in long-range LD regions) and additionally removed variants if they had > 5% missingness in any of three groups: (1) parents sequenced using the V3 exome capture kit, (2) parents sequenced using the V5 exome capture kit, or (3) controls. Samples that were outliers for differential missingness or differential MAF were excluded. PCs were generated with smartpca. The remaining samples (e.g., the DDD children) were projected onto these PCs (Supplementary Figure 1).

### Definitions of case subsets

#### Affected parents

Parents were defined as potentially “affected” if clinicians listed HPO terms for them and/or noted that they had similar phenotypes as their affected children. These analyses were only done for complete parent-child trios. In total, 1,462 trios had at least one affected parent, of which 1,246 were of inferred European genetic ancestry for use in case/control analyses and 1,306 were used for TDT analyses.

#### Diagnosed with a de novo variant

DD cases in complete parent-child trios were considered “diagnosed” with a *de novo* variant if (1) a clinician had annotated a *de novo* variant as pathogenic or likely pathogenic, (2) they were considered diagnosed with a *de novo* variant as part of an iterative analysis of the first ∼1k cases included DDD^70^, and/or (3) they had a *de novo* loss-of-function variant in a monoallelic DD-gene that had yet to be interpreted by a clinician. In total, 3,215 trios had a child with a *de novo* diagnosis, of which 2,679 were of inferred European genetic ancestry for use in the case/control analyses and 3,155 were used for TDT analyses.

#### Unaffected parents with no de novo diagnosis

These were trios that did not meet either criteria for having an affected parent or being diagnosed with a *de novo* variant. In total, 4,183 trios of inferred European genetic ancestry qualified for use in the case/control analyses and 5,124 were used for TDT analyses.

#### Environmental exposures

We used a list of 2,637 DD cases with environmental exposures as defined in Wright et al.^2^. Specifically, these DD cases were born prematurely (< 37 weeks gestation), had mothers with diabetes, and/or were exposed to antiepileptic medications in utero. Of these, we focused only on those with unaffected parents and no *de novo* diagnosis; for the case/control analysis, this was 879 individuals, and for the TDT analysis it was 1,075 individuals.

#### Affected family members

We used a list of DD cases with affected family members as defined in Wright et al. and Huang*, Wigdor* et al.^2,9^. DD cases were split in 5 categories: (1) both parents are affected and 0 or more siblings are affected (n=87); (2) one parent is affected and at least one sibling is affected (n=187); (3) one parent is affected and no siblings are affected (n=345); (4) neither parent is affected but at least one sibling is affected (n=645); (5) a non-first degree family member is affected (n=469). All numbers reported were for individuals of inferred European genetic ancestry.

### Definition of gene sets

#### Constrained

We defined constrained genes as those with a pLI (probability of being loss-of-function intolerant) score ≥ 0.9 as defined in gnomAD v2.1^26^ (n=2,699 autosomal genes).

#### DD-associated

We defined monoallelic DD-associated genes using DDG2P^28^, downloaded on June 29, 2023. Specifically, we selected genes with an allelic requirement of “monoallelic_autosomal” with either “definitive” or “strong” for the confidence category (n=666 genes).

#### Unconstrained not DD-associated

Genes that had pLI < 0.9 and were not considered monoallelic DD-associated genes were defined as unconstrained and not DD-associated (n=15,667 autosomal genes).

#### Other gene sets evaluated

Most analyses in the main text focused on all genes and the three gene lists described above. However, we also tested (1) all unconstrained genes (pLI < 0.9, n=15,911 autosomal genes); (2) all genes that were not considered monoallelic DD-associated genes (n=17,944 autosomal genes); (3) genes that were considered both constrained and monoallelic DD-associated genes (n=422 autosomal genes); and (4) genes in the top decile of MOEUF (missense observed / expected upper bound fraction, n=1,686 autosomal genes) from gnomAD v2.1^26^.

### Case versus control regressions

We included all DD cases of inferred European genetic ancestry in these analyses, including those that were not in complete parent-child trios (n=10,644 cases versus 3,943 controls). We focused on rare variants (gnomAD allele frequency ≤ 10^-5^ and allele count in the dataset < 7, equivalent to an allele frequency ∼10^-4^) and, for each individual, determined the count of alternative alleles, split by mutational consequence (e.g., loss-of-function, missense) and gene set (e.g., constrained genes, known DD genes). For all tested mutation and gene set combinations, we used a logistic regression that corrected for sex, the first 20 principal components from the inferred European genetic ancestry group only analysis, and the number of rare autosomal variants per person. The final correction has been used in recent publications^12^, and should be a conservative correction. Specifically, we ran:

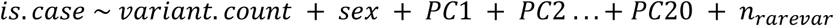

where *is.case* is 0 for controls and 1 for cases, *variant.count* is the number of rare variants in a given gene set, *sex* is the sex of the individual, *PC1* through *PC20* are the principal components, and *n_rarevar_* is the number of rare autosomal variants. Intercepts from these regressions were transformed into odds ratios using the exponential function.

See Supplemental Note for more details about modifications to these regressions, including removing the correction for the number of rare variants per person.

#### Testing significance between two regressions

To determine if the results of various regressions were different from each other (as in Supplementary Table 3), a Wald test was performed to obtain Z-scores that were transformed into p-values. Specifically,

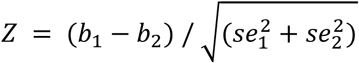

where *b_1_* is the estimate and *se_1_* is the standard error from the logistic regression for the first group, respectively, and *b_2_* is the estimate and *se_2_* is the standard error from the logistic regression for the second group, respectively.

#### Removing known de novo variants

We repeated the regressions above after removing known *de novo* variants^24^ from the trio case counts (Supplementary Figure 4). We note that this is not a fair comparison, given that the control individuals will also have a background rate of *de novo* variants, but since we do not have parental information, we do not know which variants to remove.

### Transmission disequilibrium test (TDT)

We included all DD cases, regardless of inferred genetic ancestry, that were in complete parent-child trios. For families with multiple affected children, we randomly selected one child as the representative trio for the family to avoid double counting, which resulted in 9,305 trios for analysis. We focused on rare variants (gnomAD allele frequency ≤ 10^-5^), specifically transmitted doubletons (AC=2 seen in a parent and their child) versus nontransmitted singletons (AC=1 seen only in a parent). The rate ratio reported was the number of transmitted variants to the number of nontransmitted variants with the confidence intervals determined using the rateratio.test() function in R. P-values were calculated using a ꭓ^2^ test.

#### Testing significance between two TDT rate ratios

To determine if the TDT results were different between two groups (as in Supplementary Table 6), we compared the transmitted to nontransmitted counts of each group in a 2×2 table and performed a ꭓ^2^ test to obtain a p-value.

### Other regressions

For all other regressions in this work, such as parents versus controls or male cases versus female cases, we used rare variants as defined above (e.g., gnomAD allele frequency ≤ 10^-5^ and allele count in the dataset < 7) in individuals of inferred European genetic ancestry. These were tested in a logistic regression that corrected for sex (except for sex-specific comparisons), the first 20 principal components from the inferred European genetic ancestry group only analysis, and the number of rare autosomal variants per person. We note that for the affected family member analyses, we further restricted to only unrelated individuals of inferred European ancestry to match similar work for polygenic scores^9^, but report that overall restricting to only unrelated individuals has a minimal impact on the results (Supplementary Figure 3).

### Per gene burden testing

For every gene, we tallied the number of individuals of inferred European genetic ancestry who carried a rare (gnomAD allele frequency ≤ 10^-5^ and allele count in the dataset < 7) variant in the gene, split by mutational consequence. We then performed a Fisher’s exact test to compare the rate of carriers in cases (n=10,644) versus controls (n=3,943) for pLoF variants, damaging missense (CADD ≥ 20) variants, pLoF and missense variants, and synonymous variants. We used an exome-wide significance threshold of 2.8×10^-6^.

### Estimating excess variants

To determine the excess number of variants per mutation consequence and gene list, we calculated the expected number of variants in cases by using the rate seen in controls and further corrected for the synonymous case versus control rate for the given gene list.

Specifically, this was estimated as:

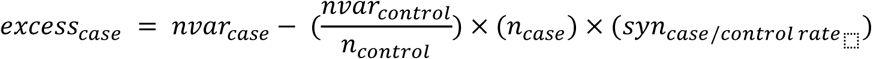

where *nvar_case_* is the number of variants in cases, *nvar_control_* is the number of variants in controls, *n_case_* is the number of cases, *n_control_* is the number of controls, and *syn_case/control rate_* is the rate of synonymous variants in cases divided by the rate of synonymous variants in controls.

### Population attributable risk

To estimate the population attributable risk (PAR), we used the following formula:

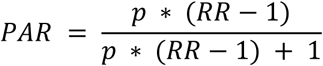

where *p* is the probability of inheriting a damaging rare variant from an unaffected parent and *RR* is the relative risk for DDs for those that inherited the given rare variants.

To estimate the *RR*, we used an estimated population prevalence of DDs of 1% and converted the odds ratio from the case/control regression of trios with unaffected parents and no diagnostic *de novo* variant for pLoF variants in constrained genes with the following formula:

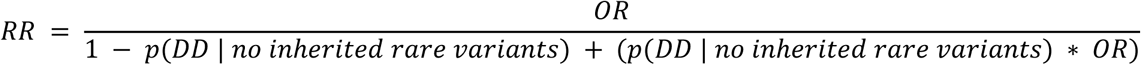

To estimate *p*, we took the rate of controls who carry a rare pLoF variant in a constrained gene (∼0.22) and used it in the following formula:

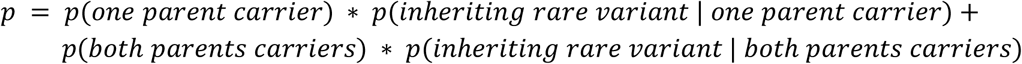

### Evaluating the s_het_ burden

In line with previous work^19^, we sought to determine a per person cumulative burden of rare variants by combining the *s_het_* selection coefficients of each gene affected by these variants via the following equation (taken from Gardner et al):

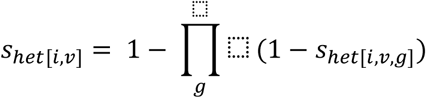

where *s_het[i,v]_* indicates individual *i*’s s_het_ burden for variant class *v* and *s_het[i,v,g]_* indicates the s_het_ score for gene *g* with a qualifying annotation for variant class *v* in individual *i*.

Here, qualifying variants were defined as:

- gnomAD allele frequency ≤ 10^-5^ and allele count in the dataset < 7
- pLoF: LOFTEE high-confidence & CADD ≥ 25
- Missense: MPC ≥ 2 & CADD ≥ 25
- Synonymous: no additional filters

Variants that fell into a gene without an *s_het_* score were not included, and individuals with no qualifying variants were given an *s_het_* burden score of 0. The specific variant filters were chosen to match what was done previously^19^. Additionally, compared to Gardner et al., we updated the selective coefficients scores to be those from Agarwal et al.^38^ (called *hs* in their work, but *s_het_* here for consistency with Gardner et al.).

We did two comparisons with these scores – within family and case versus control. For within family, we removed all known *de novo* variants^24^ to ensure that we were comparing only inherited variants. These *s_het_* burden scores were calculated for all parents and children included in the trios used for TDT analyses (n=9,305). We compared the *s_het_* burden score in children to the scores seen in parents with a Wilcoxon rank sum test. Bootstrapping with 1000 replicates was used to determine the median difference in *s_het_* burden scores as well as the 95% confidence intervals.

For the case versus control comparison, we included known *de novo* variants for the DD cases as we had no ability to remove such variants from the controls. We specifically focused on a set of 8,062 unrelated DD cases of inferred European genetic ancestry to compare to the 3,943 controls. Here, we used a probit regression of case status by scaled *s_het_* burden scores, corrected as above for sex, the first 20 principal components from the inferred European genetic ancestry group only analysis, and the number of rare autosomal variants per person.

In the regression, controls were given a weight of 1 and cases were given a weight of

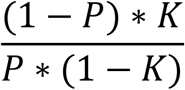

where *P* is the fraction of cases in the regression and *K* is an estimate of the population prevalence (here, 1%). This regression was also repeated to compare only unrelated DD cases with a *de novo* diagnosis (n=1,905) to controls and unrelated, undiagnosed DD cases (n=6,157) to controls.

### Determining variance explained on the liability scale

For both the *s_het_* burden scores and the rare variant counts, we wanted to determine the variance explained on the liability scale. For the *s_het_* burden scores, we used the estimate from the probit regression above and transformed it into a percent variance explained on the liability scale via the following equation:

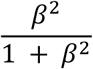

where *β* is the beta from the regression.

For the rare variant counts, we ran a probit regression with the same weights as mentioned above, replacing the scaled *s_het_* burden scores with the total counts of rare pLoF and damaging missense variants across all genes.

### Transmission and De Novo Association (TADA) test

TADA is a Bayesian framework that incorporates per-gene mutation rates, sample size, and a prior on the risk of a given variant in each gene to determine a Bayes Factor (BF) to measure the statistical evidence of association of each gene to the condition being tested. It has been used extensively to identify genes associated with autism^13,40^. We used the TADA code from Fu et al. (https://github.com/talkowski-lab/TADA_2022) to analyze our data.

We used mutation rates from gnomAD v2.1 and recalculated the mutation rate expected for missense variants with MPC ≥ 2 (“misB”) and 1 ≤ MPC < 2 (“misA”) by dividing the total missense mutation rate by the fraction of such variants possible in the gene. These missense categories matched those that were previously used^13^.

We additionally retrained the priors (or weights) used in TADA using the following data (displayed in Supplementary Figure 16):

- *De novo* analysis: *de novo* variants from 31,058 individuals with DD^24^, which includes all DDD samples used in other analyses in this work
- Case versus control analysis: rare (gnomAD allele frequency ≤ 10^-5^ and allele count in the dataset < 7) variant counts from 2,790 non-trio DD cases and from 3,943 controls of inferred European genetic ancestry
- Inherited analysis: rare (gnomAD allele frequency ≤ 10^-5^ and allele count in the dataset < 7) variants that were either transmitted from a parent to their child or not transmitted from parent to child in 9,305 trios

A comparison of the priors used in this analysis versus the Fu et al. paper is available in Supplementary Table 13, although we note that using the mutation rates and priors from the previous paper gives very similar results (Supplementary Figure 17). All results from the TADA analysis, including mutation rates, priors, Bayes Factors, and counts per gene are available in Supplementary Table 10.

