## Supplemental Note for "Substantial role of rare inherited variation in individuals with developmental disorders"

##### Correcting for the number of rare variants per person in the regressions

In the logistic regressions performed for the case/control analysis, we corrected for the number of rare autosomal variants per person, in line with prior work studying schizophrenia[11] and bipolar disorder[12]. This correction leads to a significant odds ratio (OR) < 1 for synonymous variants in cases compared to controls across all genes (OR = 0.98, p = $4x10^{-6}$, **Fig 1b**). Without this correction, we noted a significant OR > 1 for synonymous variants when comparing all European ancestry trios to controls (OR for synonymous variants in all gene = 1.03, p = $2x10^{-10}$). This elevated synonymous OR was not removed when correcting for the nonsynonymous exome-wide burden only (OR = 1.02, p = $5x10^{-5}$) or when varying the number of common variant principal components from the inferred European genetic ancestry group (ORs ~ 1.02-1.03). Additionally, we tested if this would be impacted by using canonical transcript annotations instead of the worst transcript annotation or if comparing only DD cases sequenced on the V3 or V5 baits to controls, but neither changed the ORs much. We therefore chose to use the conservative approach of correcting all logistic regressions by the number of rare autosomal variants per person.

##### Comparing rare genetic burden in parents to controls

We saw significant overtransmission of damaging genetic variants in constrained and DD-associated genes from parents, including those unaffected by similar phenotypes, to their children with developmental disorders (DDs) in the TDT analyses (e.g., **Fig 1c**). We therefore wanted to test the burden of such variation within the parents themselves. We first compared the burden of rare variants in unaffected parents (n=13,861) of inferred European genetic ancestry to controls (n=3,943) using the logistic regression described above (**Supplementary Figure 5**). We found significant differences (p < $1x10^{-3}$) in rare pLoF burden in all genes, constrained genes, and DD-associated genes as well as a significant difference in rare, damaging missense burden in constrained genes. These odds ratios (ORs) were smaller than those observed when comparing the burden in DD cases (children in complete trios, n=7,854) to controls or those observed when comparing affected parents (n=1,195) of inferred European ancestry to controls in similar regressions (**Fig 2a**).

The ORs from the affected parent regressions were also lower than those seen for the DD cases (**Fig 2a**), which is expected given that we know *de novo* variants (included here) play a large role in risk for DDs. This attenuation was more pronounced for DD-associated genes than for constrained genes. In fact, for pLoF variants in constrained genes, we found overlapping OR estimates when comparing the DD case results to those from affected parents (case OR = 1.82 [1.69-1.98] versus affected parent OR = 1.69 [1.50-1.91]).

**Investigating potential sex-based differences in burden**

The female protective effect, where females require a higher genetic burden to show traits associated with a condition, has been reported in autism[30,31] and other neurodevelopmental disorders. Along these lines, we previously reported[24] a higher rate of *de novo* nonsynonymous variants in female cases compared to male cases specifically in a set of established autosomal DD-associated genes (odds ratio = 1.16, Fisher's exact p = $4.4 \times 10^{-7}$). However, we found that the burden in autosomal genes in undiagnosed cases did not significantly differ (odds ratio = 1.03, Fisher's exact p = 0.29). We wanted to further investigate potential differences in genetic burden by sex in this study.

The most obvious way the female protective effect might present would be finding a higher burden of rare variants in female cases compared to male cases. To evaluate this, we ran the previously described case/control regressions (1) only for female cases compared to female controls (n = 1,651 and 1,940, respectively when using individuals of European genetic ancestry with unaffected parents and no *de novo* diagnosis) and (2) only for male cases compared to male controls (n = 2,532 and 2,003, respectively when using individuals of European genetic ancestry with unaffected parents and no *de novo* diagnosis). There is a slightly higher odds ratio (OR) estimate for rare pLoF variants in DD-associated genes (**Fig 3a**), but the difference was not significant. Similarly, we found similar estimates when using only female cases or only male cases in the previously described TDT analysis (n = 2,017 female and 3,107 male cases of all inferred genetic ancestries but with unaffected parents and no *de novo* diagnosis; **Supplementary Figure 8**).

When comparing the rare variant burden in unaffected parents to controls, we found a slightly higher OR of rare, damaging variants in mothers compared to female controls versus fathers compared to male controls for pLoF variants in DD-associated genes (mother OR = 1.44 and p = $5.9 \times 10^{-4}$, father OR = 1.05 and p = 0.643; **Fig 2b**) with a more attenuated signal in constrained genes (**Supplementary Figure 6**). To test if these differences were significant, we performed a logistic regression as above comparing unaffected mothers to unaffected fathers but found nothing significant (**Supplementary Figure 7**).

Finally, we tested if the burden of rare variants inherited from mothers differed from that inherited from fathers. For this, we used a set of 6,087 DD cases from complete trios where both parents were considered unaffected and all three individuals were of inferred European genetic ancestry. We used a $\chi^2$ test to evaluate any significant differences, but found none (**Supplementary Table 7**).

**Details of the population attributable risk calculation**

We were interested in calculating the population attributable risk (PAR) for rare inherited pLoF variants in constrained genes, a set that showed enrichment in multiple analyses. Said another way, if unaffected parents never transmitted these rare pLoF variants in constrained genes, what fraction of DD cases would be eliminated?

This PAR calculation required a few estimates. First, we needed to use an odds ratio (OR) from the case/control regression to get a relative risk of DDs for those who inherited a rare pLoF in a constrained gene. If we use the OR from the case/control regression of trios with unaffected parents and no *de novo* diagnosis (OR = 1.49), we estimated the PAR to be 8.37%. However, this OR includes *de novo* variants. When using the OR from the case/control regression of the same set of trios when known *de novo* variants were removed (OR = 1.34), the estimated PAR dropped to 5.94%. As mentioned above, the OR from the analysis with *de novo* variants removed is likely an underestimate since we were unable to remove *de novo* variants from controls.

Second, we needed to use an estimate of the population prevalence of DDs when calculating the relative risk. Here, we used 1% in line with other recent work[9]. However, if we varied the population prevalence estimate between 0.2% and 1.5%, we found that it had a minimal impact on the PAR estimate: the PAR estimate ranged from 8.31-8.46% versus 8.37% when using 1% as the population prevalence.

Finally, we made a few simplifying and conservative assumptions. Given that we are studying a rare condition, we used the ORs as an approximation of the relative risk. For our calculation of the probability of inheriting a rare damaging variant from an unaffected parent, we assumed no assortment on rare damaging variants and ignored different effects for inheriting one versus two or more of these rare variants, both of which should be conservative assumptions.

**Repeating TADA with autism-based mutation rates and priors**

The TADA framework requires providing mutation rates per gene and priors for multiple classes of genetic variants (e.g., *de novo* pLoF, inherited missense with MPC ≥ 2, etc). In Fu et al.[13], they used gnomAD v2.1 based mutation rates but subsequently modified them based on the rate of *de novo* variants seen in the neurotypical siblings sequenced as part of their study. As we had no siblings here to modify rates, we chose to use the provided gnomAD v2.1 rates. Additionally, given the different genetic architectures of developmental disorders (DDs) compared to autism – mostly notably the greater contribution of *de novo* variation to DDs – we retrained the priors used in TADA based on the data available in this study (**Supplementary Figure 16**; a comparison of priors is listed in **Supplementary Table 13**). We used these updated priors in the main text.

We also wanted to evaluate how the results would look when using the mutation rates and priors from the autism analysis. We found a strong correlation between the results of TADA when using our remade DDD-based mutation rates and priors compared to results when using the provided mutation rates and priors from the Fu et al. paper (Pearson correlation coefficient =

0.833; **Supplementary Figure 17**). There were a similar number of genes that cross the exome-wide significance threshold ($p < 2.8x10^{-6}$): 269 with DDD-based rates and priors and 282 with the autism-based rates and priors, nearly all of which (n=257; 91-95%) are significant in both analyses. Of the 26 seen uniquely in the autism-based analysis, 23 were significant in our prior study of *de novo* variants in 31,058 DD trios[24] or were known DD-associated genes.

##### Breakdown of Bayes Factors from the TADA analysis

The TADA framework generates Bayes Factors (BF) for every class of genetic variants tested (e.g., *de novo* pLoF) per gene as a measure of the strength of evidence for that class of variants. We summed these BFs both by mutational class (e.g., pLoF) and inheritance type (e.g., *de novo*) for the 269 genes that achieved significance ($p < 2.8x10^{-6}$). We found that pLoF BF made up the majority of the contribution for mutational class (66.7%; **Fig 4a**; **Supplementary Table 11a**), which matches what prior autism studies have reported. More noticeable was that nearly all of the BF signal for these 269 genes came from *de novo* variants (97.2%; **Fig 4b**; **Supplementary Table 11b**).

We were further interested in whether these BF contributions varied by whether the genes were previously known DD-associated genes (n=244) or genes with more limited evidence of DD-association (n=25). We found that the 25 genes with more limited evidence of DD-association have a stronger contribution from pLoF variants (76% of BF contribution versus 66.5% in the known genes; **Fig 4d**). More strikingly, for inheritance class BFs, the new genes have a greater contribution from both case/control (9.7% of BF contribution versus 2.4% in the known genes) and inherited (1.5% versus 0.3%) variants (**Fig 4e**). Comparisons of BFs are listed in **Supplementary Table 11**, including to the ~16k genes that were not significant in the TADA analysis.

##### Classification of genes

For the 25 TADA significant genes that were not either significant in our prior analysis of *de novo* variation[24] or considered definitive or strong monoallelic DD-associated genes via DDG2P[28] as of July 2023, we wanted to establish how likely they were to be true DD-associated genes. To do this, we collected information from a number of sources, including their p-values in the prior *de novo* study including in both the full and undiagnosed analyses[24], five scores of selective constraint (pLI[26], LOEUF[26], $s_{het}$[44], pHaplo[45], and pTriplo[45]), gene-phenotype databases (DDG2P[28], OMIM[41], GenCC[42], PanelApp[43]), and literature searches. Our approximate decision tree for classifying the genes is depicted in **Supplementary Figure 14** with a visual summary of the evidence in **Supplementary Figure 15**.

The 11 genes we classified as having a high likelihood of being true DD-associated genes either were listed in gene-phenotype databases with ties to DD-related phenotypes (e.g., *SYNCRIP* is listed as Green in PanelApp for intellectual disability) and/or had a recent publication describing a cohort of individuals with mutations in these genes (e.g., *FOSL2*[46] and *ZFHX3*[71]).

The 13 genes we classified as having a medium likelihood of being true DD-associated genes were primarily driven by strong selective constraint scores and/or having some prior association to autism (frequently via FDR-based methods). However, all genes were kept in this category unless there was a reason to move them to the low likelihood group (see below). This meant that the known biallelic DD-associated gene, *SLC39A8*, is included on this medium list even though it does not have a low LOEUF score or any prior ties to autism.

Only one gene is listed as a low likelihood of being DD-associated. *GLYR1*'s significance in TADA was primarily from *de novo* pLoF variants (BF = 927). However, two of the three individuals with a *de novo* pLoF in *GLYR1* had an additional nonsynonymous *de novo* variant in a known DD-associated gene or one that was significant in our prior analysis[24] and were considered “diagnosed” in that work. The third individual is from DDD and also has a nonsynonymous *de novo* in another gene, which has been classified as “uncertain” by the clinical team. There was some signal for *de novo* missense variants with MPC ≥ 2 (misB BF = 9.25) and for variants in the case/control analysis (BF = 1.76, 3.52, 1.45 for pLoF, misB, and misA, respectively), but these lines of evidence alone likely wouldn't be enough for the gene to cross the significance threshold without the *de novo* pLoF contribution. Of note, we have found that 3-5% of DDD cases have a dual molecular diagnosis, so we cannot completely rule out this gene as a candidate[2].

**Comparing the rate of diagnosed cases for variant carriers in the genes with a high likelihood of being true DD genes**

There were eleven genes that we classified as having a high likelihood of being a true DD-associated gene based on recent publications or inclusion in diagnostic gene lists (i.e., *BAP1*, *FOSL2*, *KCNT2*, *PSMC3*, *RFX7*, *SEMA6B*, *SRSF1*, *SYNCRIP*, *VCP*, *ZBTB7A*, *ZFHX3*). We hypothesized that, if these were true DD genes, we may find that individuals carrying damaging genetic variants in these genes would have a lower rate of being diagnosed compared to the overall rate we find in our cohort. For individuals harboring damaging genetic variants in these eleven genes, we checked if they were considered diagnosed by an alternative variant.

*Defining the set of diagnosed individuals*

For DDD individuals, we had access to clinician classifications of reported genetic variants via DECIPHER[72]. We defined DDD individuals as diagnosed if they had a reported diagnostic variant that was classified as pathogenic or likely pathogenic by the clinical teams responsible for the individual. We previously reported that ~33% (4,484/13,449) of individuals in DDD have a clinician classified diagnostic variant[2].

For non-DDD individuals included in the *de novo* analysis, we did not have access to full diagnostic information. We therefore defined individuals as “diagnosed” if they had a nonsynonymous (i.e., pLoF or missense) *de novo* variant in a significant or well-established DD-associated gene, in line with our approach in Kaplanis*, Samocha*, Wiel*, Zhang* et al.[24]. In total, this represented 5,044 of the 21,200 non-DDD individuals (diagnostic rate of ~24%).

*Defining the set of individuals harboring damaging genetic variants*

Given that some classes of genetic variants were not contributing to the significance of a gene in TADA, we wanted to use a BF threshold to define the variants that were contributing to the signal and thereby only evaluate individuals harboring those specific classes of variants. For example, *ZFHX3* was driven solely by *de novo* pLoF variants (BF of $1.1 \times 10^{10}$ with all other BF ≤ 1) and we therefore only evaluated the 11 individuals with *de novo* pLoF variants in this gene. We set the BF threshold at 1.3, since it included contributions from *de novo*, case/control, and inherited analyses. At this threshold, 12 of the 85 individuals with damaging genetic variants had an alternative diagnosis (14.1%).

*Comparing the rate of diagnoses*

We made a composite diagnostic rate between DDD and non-DDD individuals for our comparator by adding diagnosed individuals and dividing by the total number considered. This worked out to be 9,528 (4,484 + 5,044) diagnosed individuals out of 34,649 (13,449 + 21,200) total individuals, giving a diagnostic rate of 27.5%.

We found that the diagnostic rate of individuals with damaging genetic variants in one of the 11 high likelihood DD genes was significantly lower than the overall diagnostic rate in DDD (14.1% versus 27.5%, Fisher's exact $p = 4.8 \times 10^{-3}$). However, if we were to use a much lower BF threshold (>1), the diagnostic rate of 22.5% (25/111 individuals) is not significantly lower than the 27.5% overall diagnostic rate (Fisher's exact p = 0.287).

#### Removing affected parents from the analysis of trios with a diagnostic *de novo* variant

In concurrent work, we reported that DD cases with a monogenic diagnosis–encompassing *de novo*, recessive, dominant, and X-linked diagnoses–have a higher common polygenic risk from controls, but that this signal was driven entirely by the affected parents in the diagnosed cases[9]. In this work, we found that individuals with a likely diagnostic *de novo* variant had a significant burden of rare pLoF and damaging missense variants in a case/control regression. Notably, this signal remained significant for pLoFs in constrained and DD-associated genes, even when removing the known *de novo* variants[24] (**Supplementary Figure 4**). To evaluate if this was also driven by affected parents, we repeated the case/control analysis for the trios with a diagnostic *de novo* variant after removing those trios with one or more affected parents (n=2,425 DD cases remaining). We found very similar results when comparing to the full set of trios with a *de novo* diagnosis (n=2,679), even when removing *de novo* variants (**Supplementary Table 12**).

### Supplementary Figures

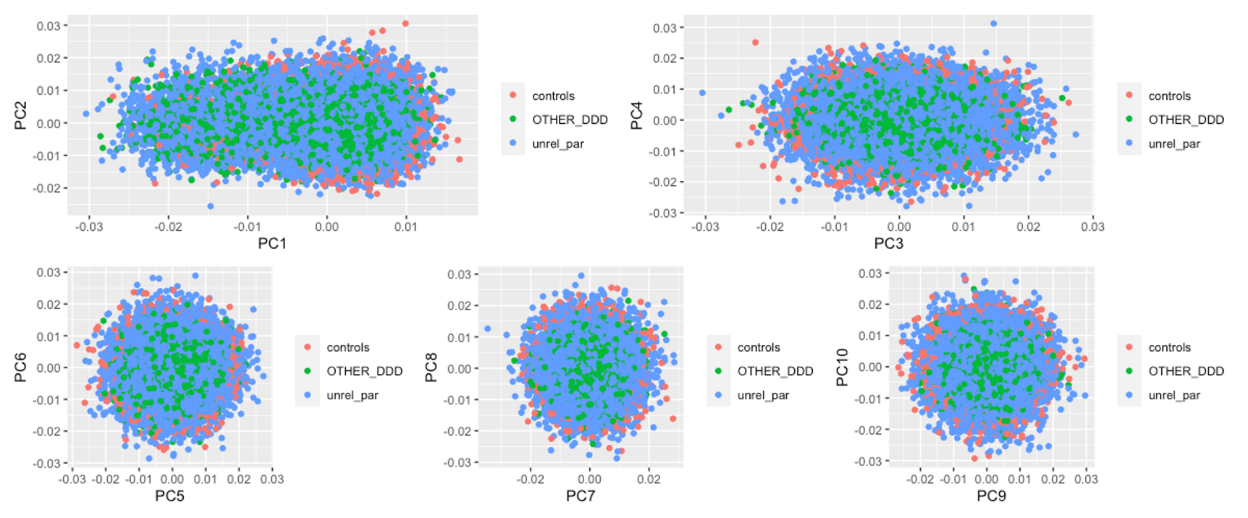


**Supplementary Figure 1**. Comparison of principal components (PCs) for the set of 32,065 individuals from the DDD study and 3,943 individuals from the INTERVAL study with inferred European genetic ancestry. The unrelated parents (in blue) and controls (in red) were used to generate these within-European ancestry PCs. The other DDD samples (primarily children, but some related parents; in green) were projected onto these PCs.

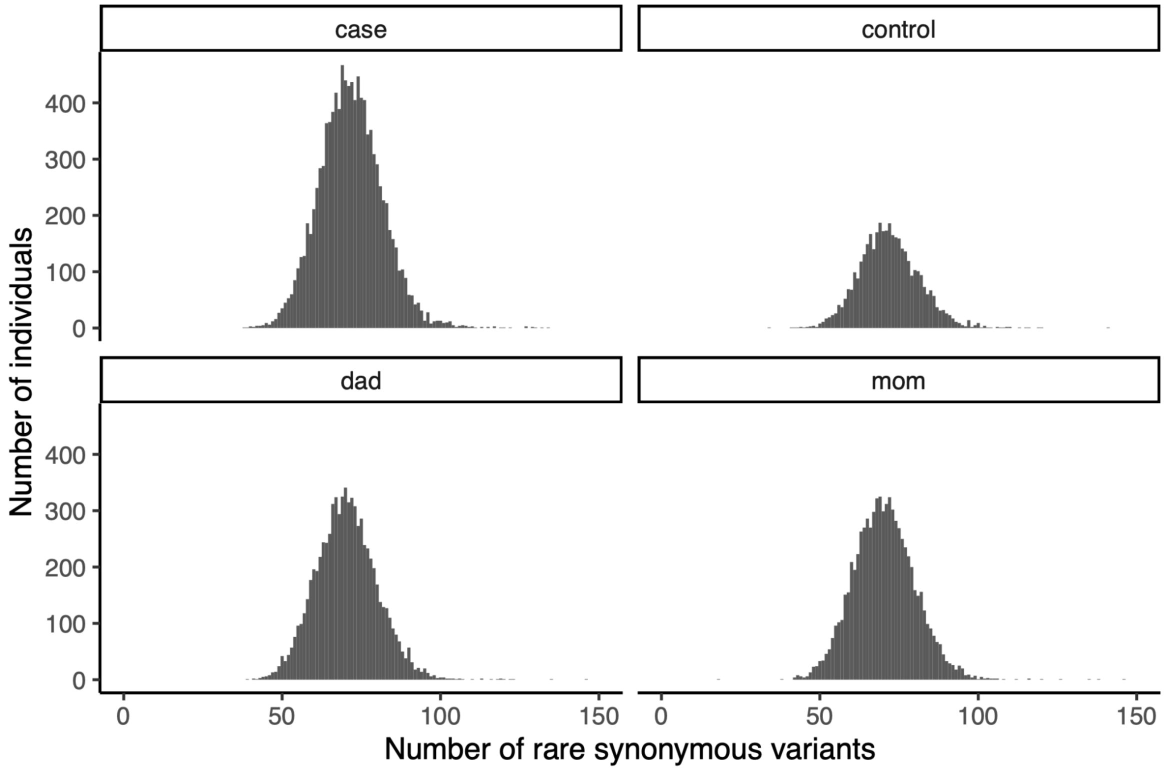


**Supplementary Figure 2**. Number of rare (allele frequency < 0.001) synonymous variants in European genetic ancestry developmental disorder cases (n=10,644), their mothers (n=7,530), their fathers (n=7,527), and controls (n=3,943). X-axis is limited to 150; there were five DD cases, six fathers, and one mother with rare synonymous counts >150.

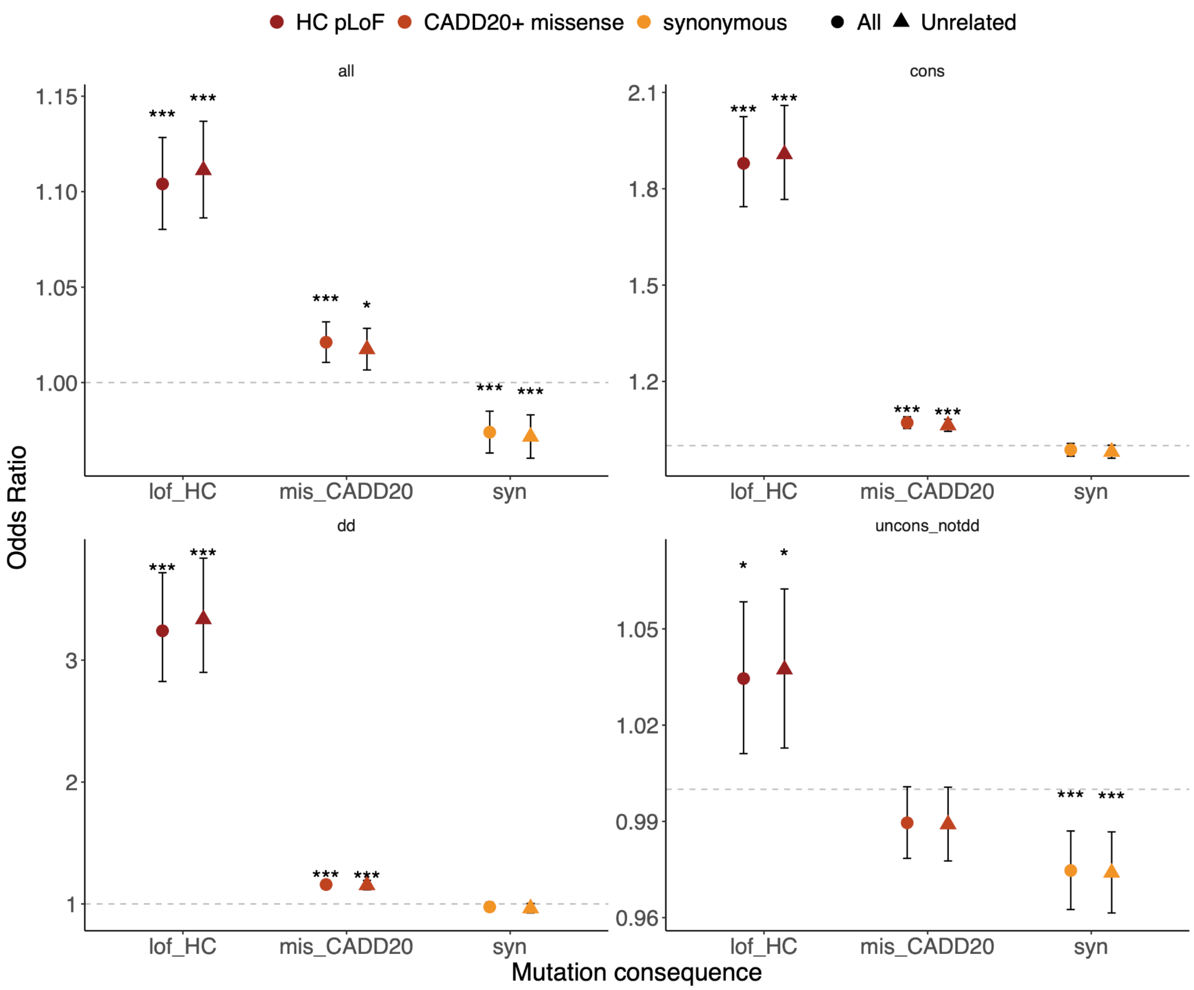


**Supplementary Figure 3**. Rare variant burden odds ratios from a case versus control regression for European genetic ancestry individuals. In circles, all developmental disorder cases (n=10,644) are compared to controls (n=3,943). In squares, unrelated cases (n=8,415) are compared to controls. LOFTEE high-confidence predicted loss-of-function (HC pLoFs) are shown in dark red; missense variants with CADD ≥20 in orange; and synonymous variants in yellow. Four gene sets shown: all genes ("all", n=18,610, top left), constrained ("cons", pLI ≥0.9, n=2,699, top right), monoallelic DD-associated ("dd", n=666, bottom left), and unconstrained genes with no prior monoallelic DD-association ("ucons_notdd", n=15,667, bottom right). * $1x10^{-3} \leq p < 1x10^{-2}$; ** $1x10^{-4} \leq p < 1x10^{-3}$; *** $p < 1x10^{-4}$.

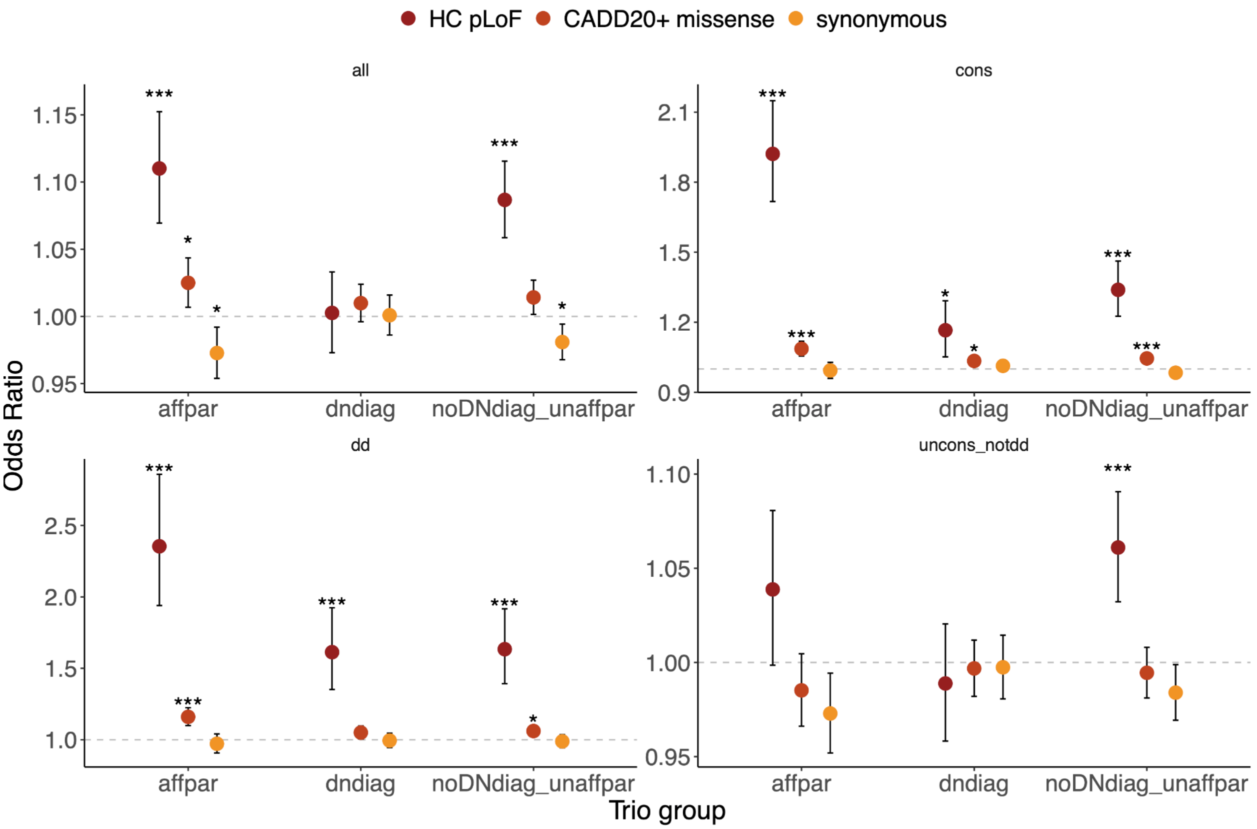


**Supplementary Figure 4**. Rare variant odds ratios from a case versus control regression for individuals of European genetic ancestry when removing known *de novo* variants from the case counts. Three trio groups are shown: developmental disorder cases in full trios with at least one affected parent (“affpar”, n=1,246), cases in full trios who have a *de novo* diagnosis (“dndiag”, n=2,679), and cases in full trios with unaffected parents and no *de novo* diagnosis (“noDNdiag_unaffpar”, n = 4,183). Control sample size is 3,943. LOFTEE high-confidence predicted loss-of-function (HC pLoFs) are shown in dark red; missense variants with CADD ≥20 in orange; and synonymous variants in yellow. Four gene sets shown: all genes (“all”, n=18,610, top left), constrained (“cons”, pLI ≥0.9, n=2,699, top right), monoallelic DD-associated (“dd”, n=666, bottom left), and unconstrained genes with no prior monoallelic DD-association (“ucons_notdd”, n=15,667, bottom right). * $1x10^{-3} \leq p < 1x10^{-2}$; ** $1x10^{-4} \leq p < 1x10^{-3}$; *** $p < 1x10^{-4}$.

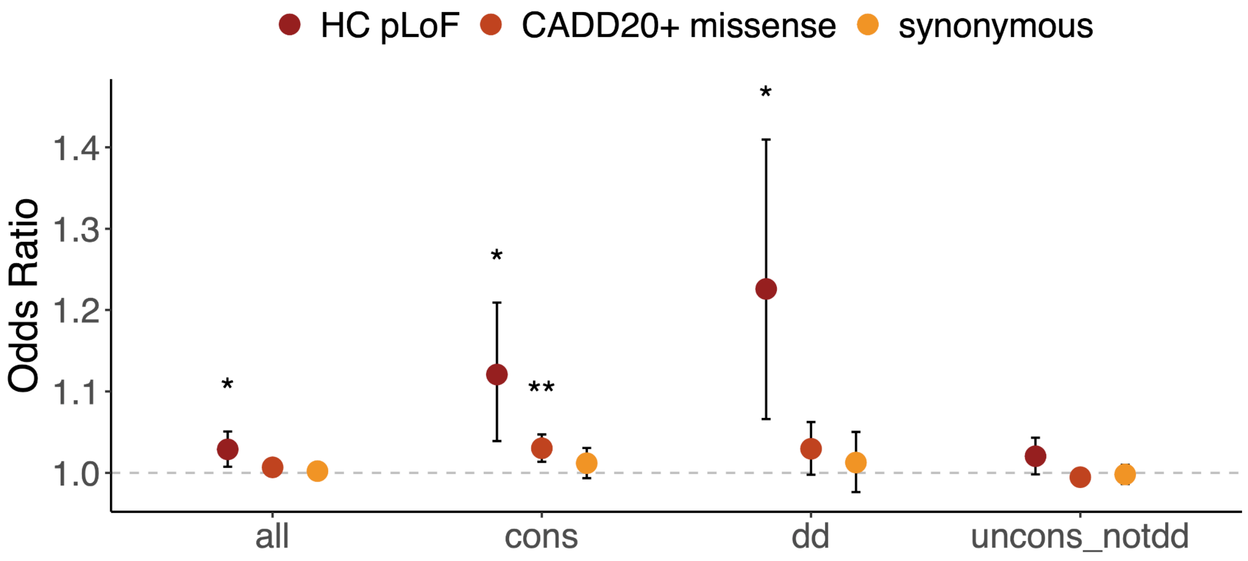


**Supplementary Figure 5**. Odds ratios when comparing rare variant burdens in 13,861 unaffected parents to 3,943 ancestry-matched controls. LOFTEE high-confidence predicted loss-of-function (HC pLoFs) are shown in dark red; missense variants with CADD ≥20 in orange; and synonymous variants in yellow. Four gene sets shown: all genes (“all”, n=18,610), constrained (“cons”, pLI ≥0.9, n=2,699), monoallelic DD-associated (“dd”, n=666), and unconstrained genes with no prior monoallelic DD-association (“ucons_notdd”, n=15,667). * $1x10^{-3} \leq p < 1x10^{-2}$; ** $1x10^{-4} \leq p < 1x10^{-3}$; *** $p < 1x10^{-4}$.

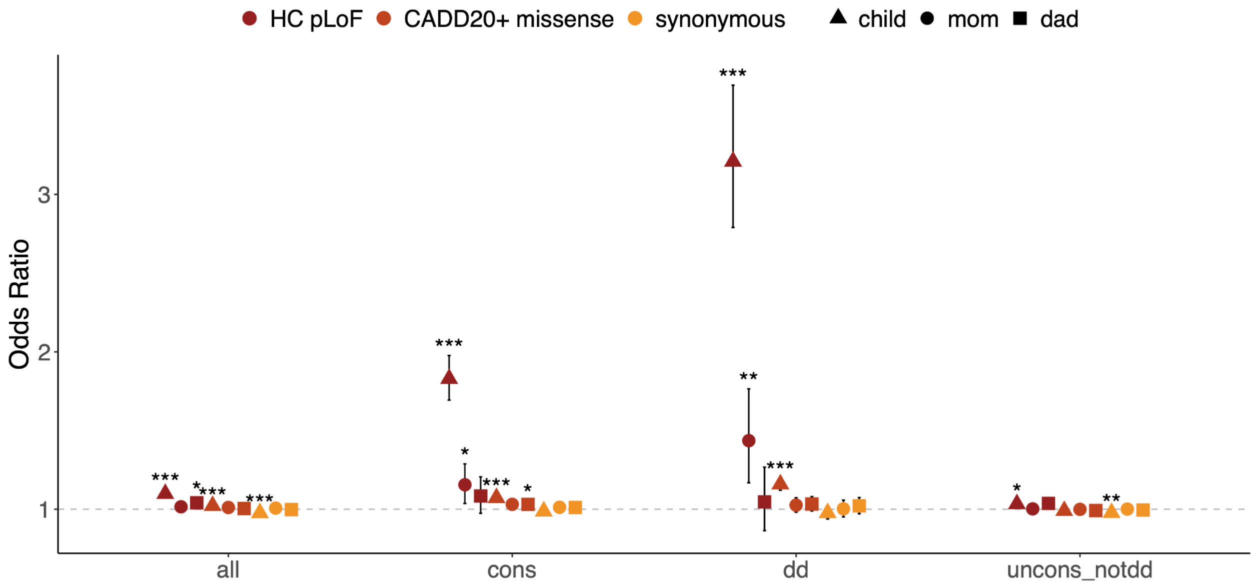


**Supplementary Figure 6**. Comparing the odds ratios of rare variant burden regressions compared to controls for individuals of European genetic ancestry: developmental disorder cases in complete trios (“child”, triangles, n=7,854), unaffected mothers (“mom”, circles, n=6,872), and unaffected fathers (“dad”, squares, n=6,989). LOFTEE high-confidence predicted loss-of-function (HC pLoFs) are shown in dark red; missense variants with CADD ≥20 in orange;

and synonymous variants in yellow. Four gene sets shown: all genes (“all”, n=18,610), constrained (“cons”, pLI ≥0.9, n=2,699), monoallelic DD-associated (“dd”, n=666), and unconstrained genes with no prior monoallelic DD-association (“ucons_notdd”, n=15,667). * $1x10^{-3} \leq p < 1x10^{-2}$; ** $1x10^{-4} \leq p < 1x10^{-3}$; *** $p < 1x10^{-4}$.

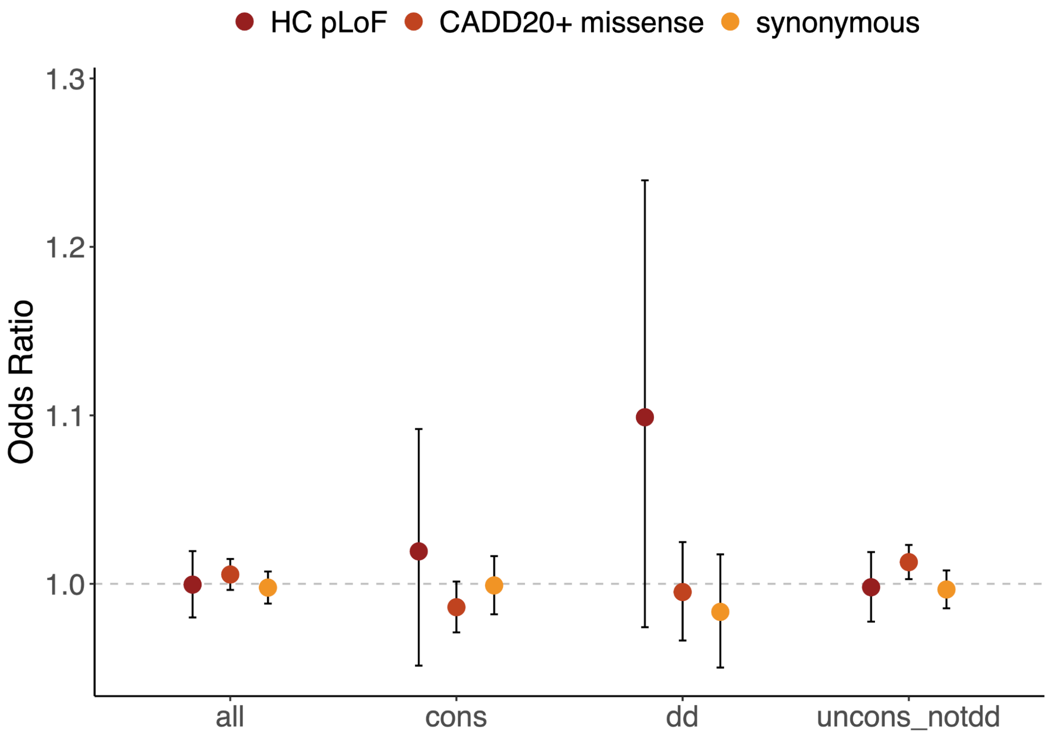


**Supplementary Figure 7**. The odds ratios for rare variant burden from a mother versus father regression for European genetic ancestry parents that are unaffected (n = 6,872 mothers and 6,989 fathers). LOFTEE high-confidence predicted loss-of-function (HC pLoFs) are shown in dark red; missense variants with CADD ≥20 in orange; and synonymous variants in yellow. Four gene sets shown: all genes (“all”, n=18,610), constrained (“cons”, pLI ≥0.9, n=2,699), monoallelic DD-associated (“dd”, n=666), and unconstrained genes with no prior monoallelic DD-association (“ucons_notdd”, n=15,667).

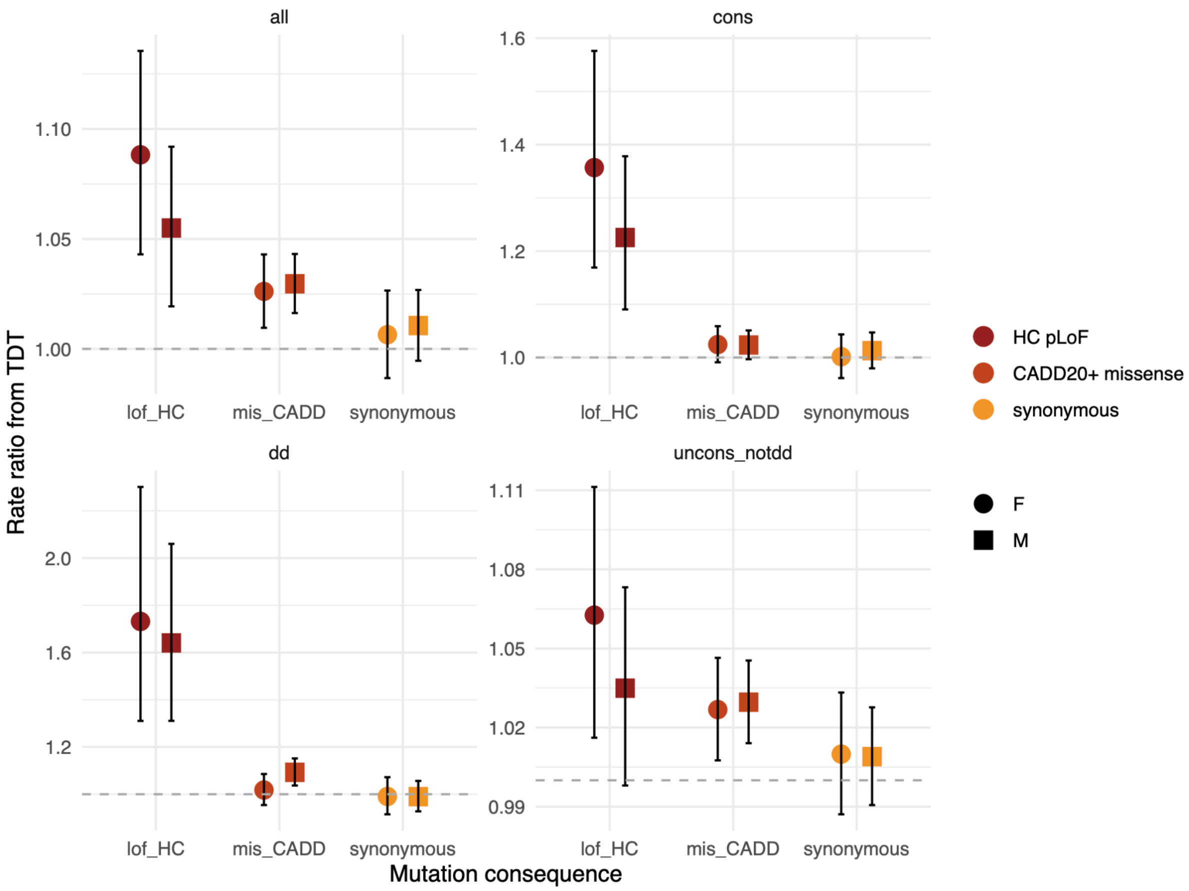


**Supplementary Figure 8.** Rate ratios from the transmission disequilibrium test (TDT) when evaluating only female cases (“F”, circles, n=2,017) or only male cases (“M”, squares, n=3,107). All genetic ancestries are used for this analysis, but are limited to only those cases with unaffected parents and no *de novo* diagnosis. LOFTEE high-confidence predicted loss-of-function (pLoFs) are shown in dark red; missense variants with CADD ≥20 in orange; and synonymous variants in yellow. Four gene sets shown: all genes (“all”, n=18,610, top left), constrained (“cons”, pLI ≥0.9, n=2,699, top right), monoallelic DD-associated (“dd”, n=666, bottom left), and unconstrained genes with no prior monoallelic DD-association (“ucons_notdd”, n=15,667, bottom right).

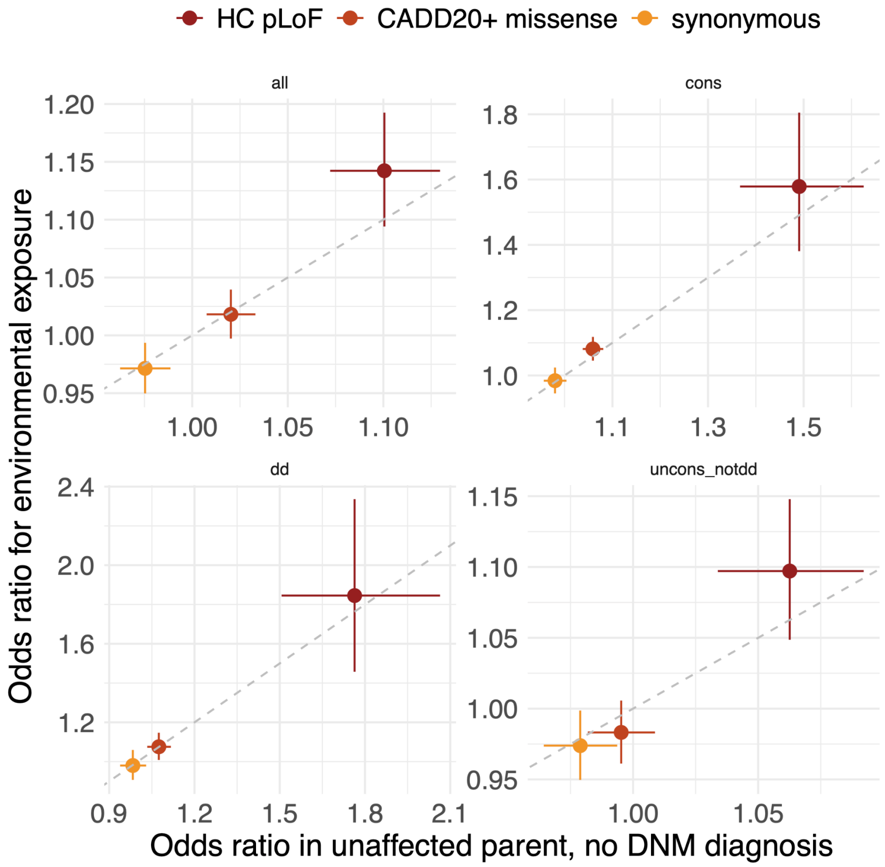


**Supplementary Figure 9**. The odds ratios from case versus control regressions for European genetic ancestry developmental disorder cases with unaffected parents and no *de novo* diagnosis (n = 4,183, x-axis) compared to that set of cases who also had an environmental exposure (i.e., premature birth, maternal diabetes, and/or exposure to antiepileptic medications in utero; n = 879, y-axis). LOFTEE high-confidence predicted loss-of-function (HC pLoFs) are shown in dark red; missense variants with CADD ≥20 in orange; and synonymous variants in yellow. Four gene sets shown: all genes (“all”, n=18,610, top left), constrained (“cons”, pLI ≥0.9, n=2,699, top right), monoallelic DD-associated (“dd”, n=666, bottom left), and unconstrained genes with no prior monoallelic DD-association (“ucons_notdd”, n=15,667, bottom right).

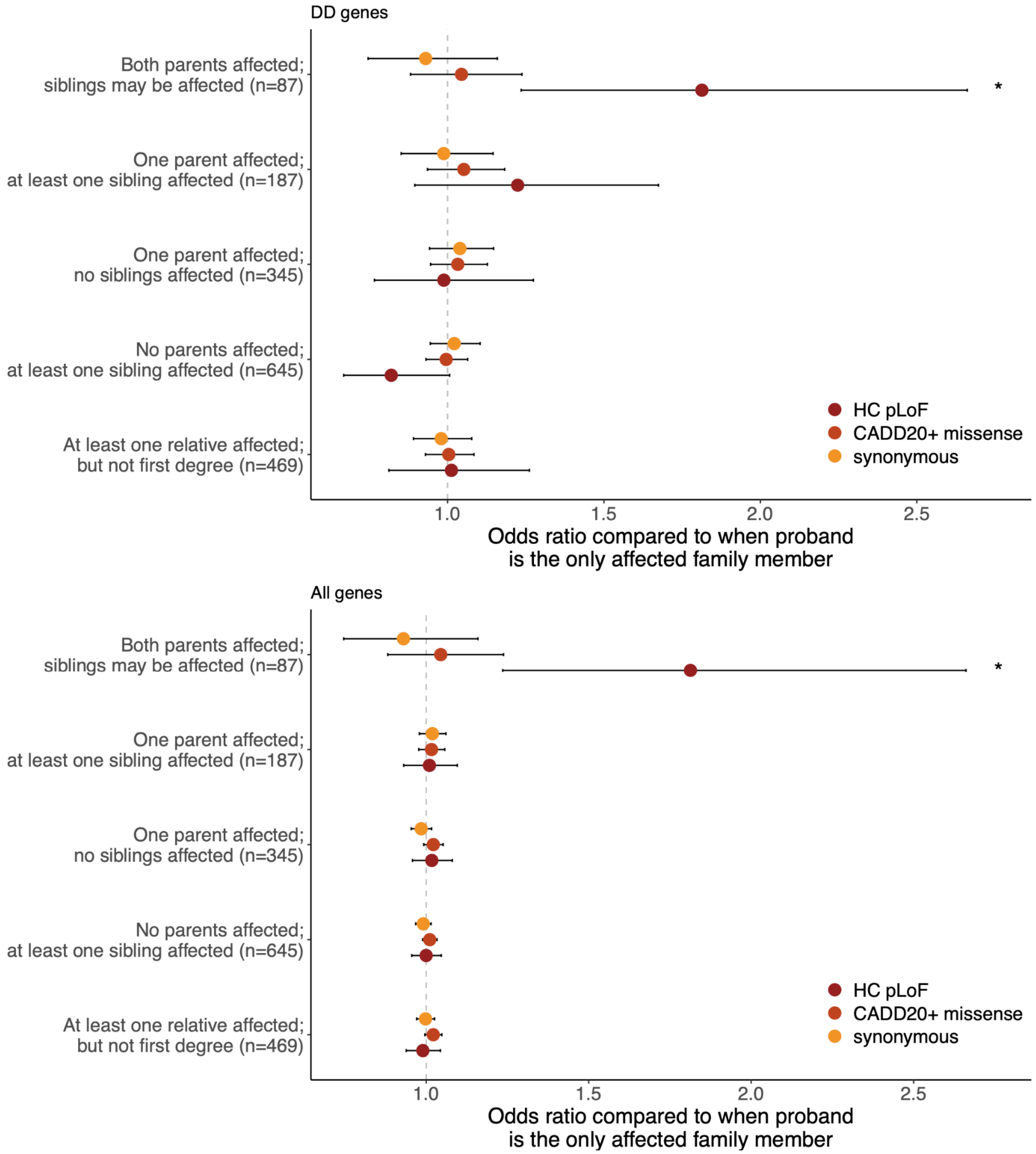


**Supplementary Figure 10**. Odds ratios in monoallelic developmental disorder (DD)-associated (top panel) or all genes (bottom panel) from a regression comparing unrelated European genetic ancestry cases with and without additional affected family members, split by the number and type of relative affected. All comparisons are against the burden in cases that are the only affected family member (n = 6,664). LOFTEE high-confidence predicted loss-of-function (HC pLoFs) are shown in dark red; missense variants with CADD ≥ 20 in orange; and synonymous variants in yellow. * $1x10^{-3} \leq p < 1x10^{-2}$; ** $1x10^{-4} \leq p < 1x10^{-3}$; *** $p < 1x10^{-4}$.

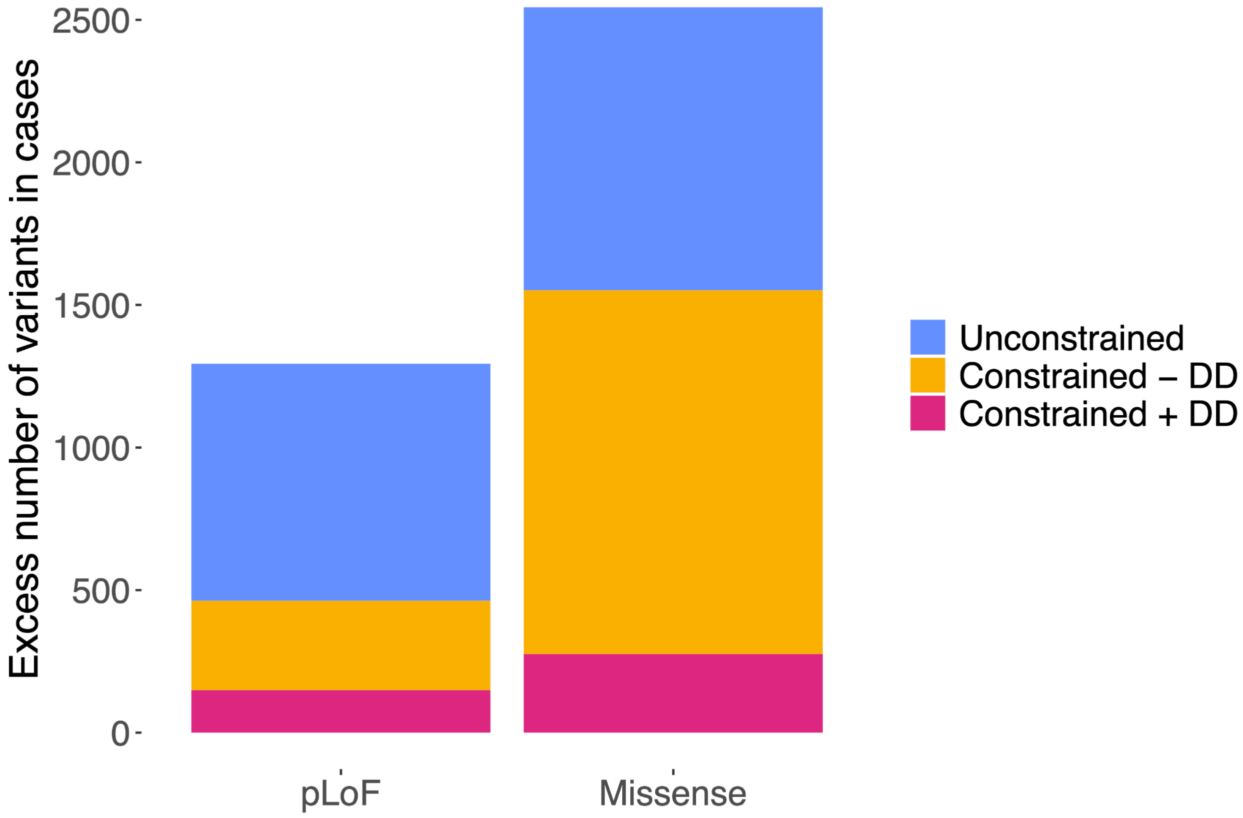


**Supplementary Figure 11.** Estimated excess of rare damaging variants in developmental disorder (DD) cases compared to controls. For each gene set, we estimated the excess of variants split by mutational consequence (e.g., loss-of-function, missense) by using the rate of such variants in controls multiplied by the number of DD cases and corrected for the ratio of rate of rare synonymous variants found in cases compared to the rate seen in controls. For this plot, we compared the 4,183 European genetic ancestry DD cases with unaffected parents and no *de novo* diagnosis to 3,943 ancestry-matched controls. Shown are the excesses for LOFTEE high-confidence predicted loss-of-function (pLoF) and missense variants with CADD ≥ 20 in three gene sets: constrained (pLI ≥ 0.9) genes with a prior monogenic DD-association (n=422; pink), constrained genes without a prior monogenic DD-association (n=2,277; yellow), and unconstrained genes with no DD-association (n=15,667; blue).

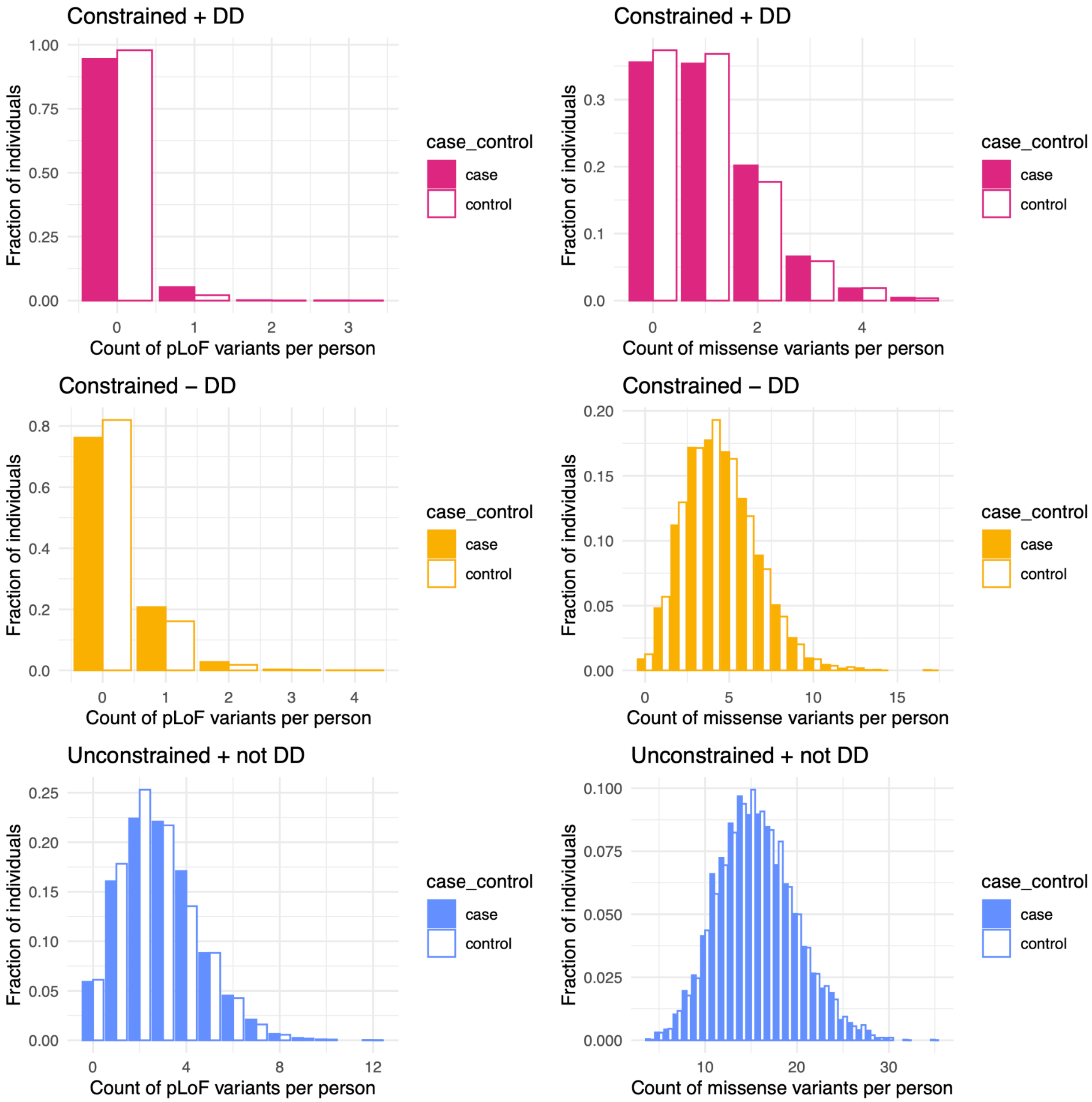


**Supplementary Figure 12**. Counts of loss-of-function and damaging missense variants per person comparing developmental disorder (DD) cases and ancestry-matched controls. For LOFTEE high-confidence predicted loss-of-function (pLoF) variants (left panel) and missense variants with CADD ≥20 (right panel), we calculated the number of rare variants per person in a set of 4,183 European genetic ancestry DD cases with unaffected parents and no *de novo* diagnosis (filled bars) and 3,943 ancestry-matched controls (white bars). These values are calculated for three gene sets: constrained (pLI ≥ 0.9) genes with a prior monogenic DD-association (n=422; pink), constrained genes without a prior monogenic DD-association (n=2,277; yellow), and unconstrained genes with no DD-association (n=15,667; blue).

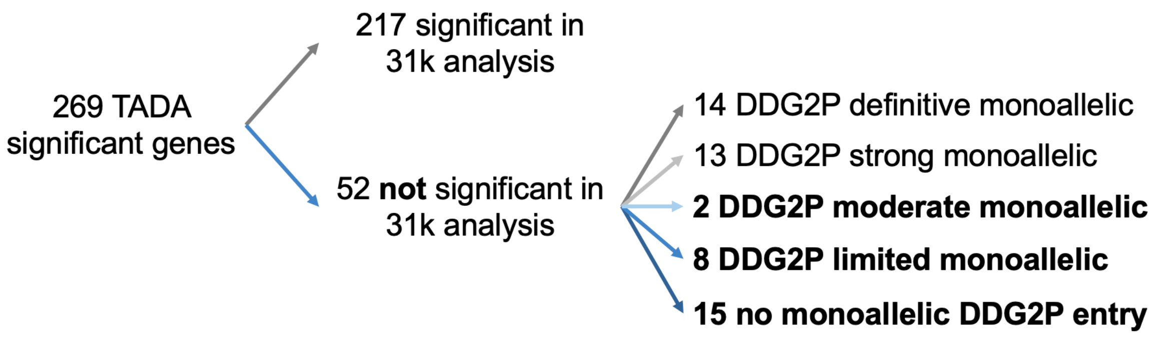


**Supplementary Figure 13**. There were 269 genes significant ($p < 2.8x10^{-6}$) in the TADA analysis. These were further filtered to remove those 217 genes that were significant in the prior study of *de novo* variants ("31k analysis"), and then split based on their entries (or lack thereof) in a curated developmental disorder associated gene list (DDG2P) as of July 2023. The focus here was specifically on those genes with known monoallelic associations. This left 25 genes (bold); 2 with moderate monoallelic association to developmental disorders, 8 with limited monoallelic association to developmental disorders, and 15 with no monoallelic association to developmental disorders.

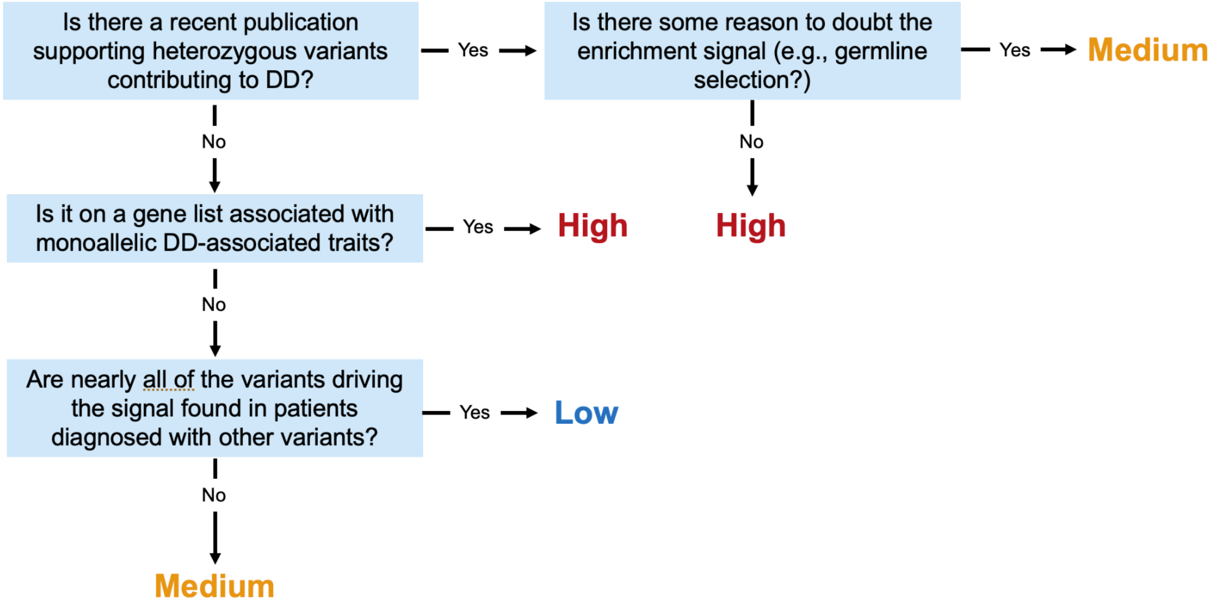


**Supplementary Figure 14**. Approximate decision tree for classifying the 25 TADA significant genes with limited to no prior developmental disorder (DD) associations into "high", "medium", and "low" likelihoods of being *bona fide* DD-associated genes.

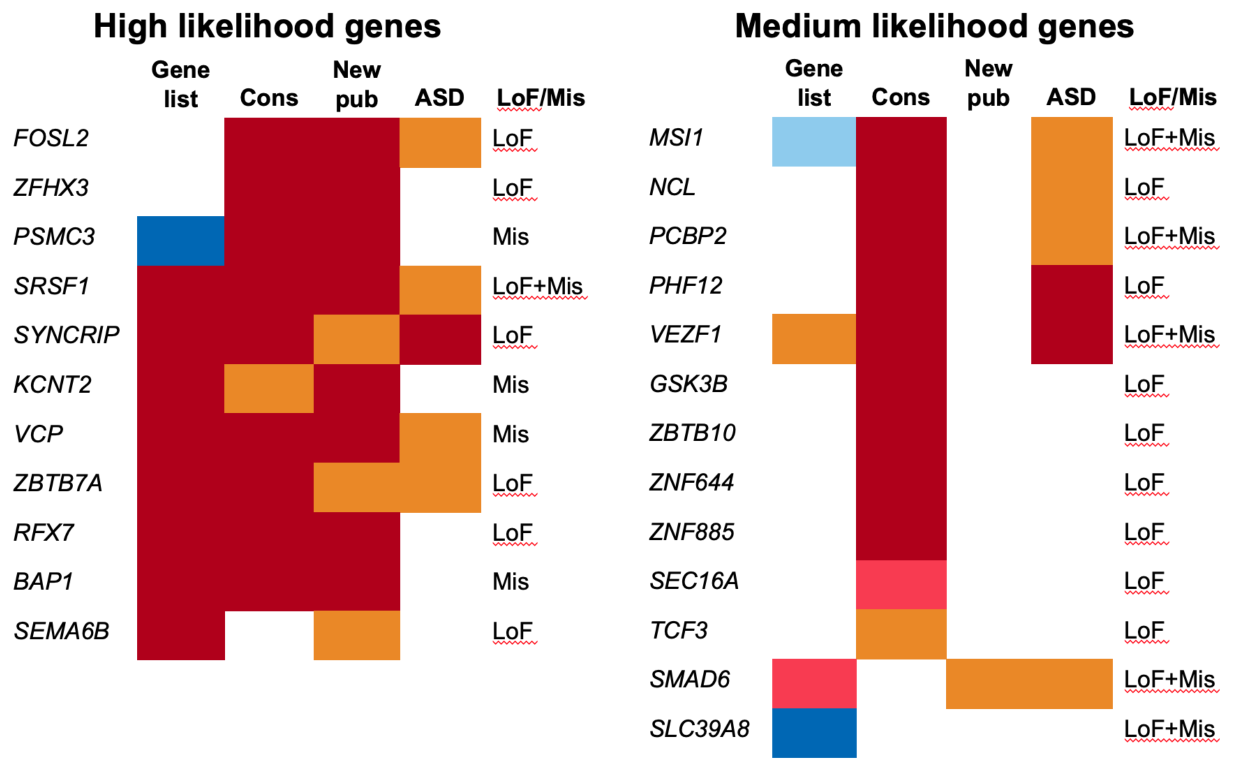


**Supplementary Figure 15**. Visual summary of the evidence to classify the 25 TADA significant genes with limited to no prior developmental disorder (DD) associations split by high versus medium likelihood of being a DD-associated gene. One low likelihood gene, *GLYR1*, is not shown. Evidence includes:

(1) “gene list” – presence on a non-DDG2P gene list such as PanelApp (red for monoallelic, blue for biallelic)
(2) “cons” – constraint scores such as pLI, LOEUF, pHaplo, and pTriplo (darker red means more constrained)
(3) “new pub” – more recent publication with some evidence of association to DDs (darker red = publication; orange = associated to autism)
(4) “ASD” – prior association to autism, typically via FDR-based frameworks (orange for some association, dark red for Bonferroni-corrected association)
(5) “LoF/Mis” – whether the TADA signal was driven by loss-of-function (LoF) and/or missense variants

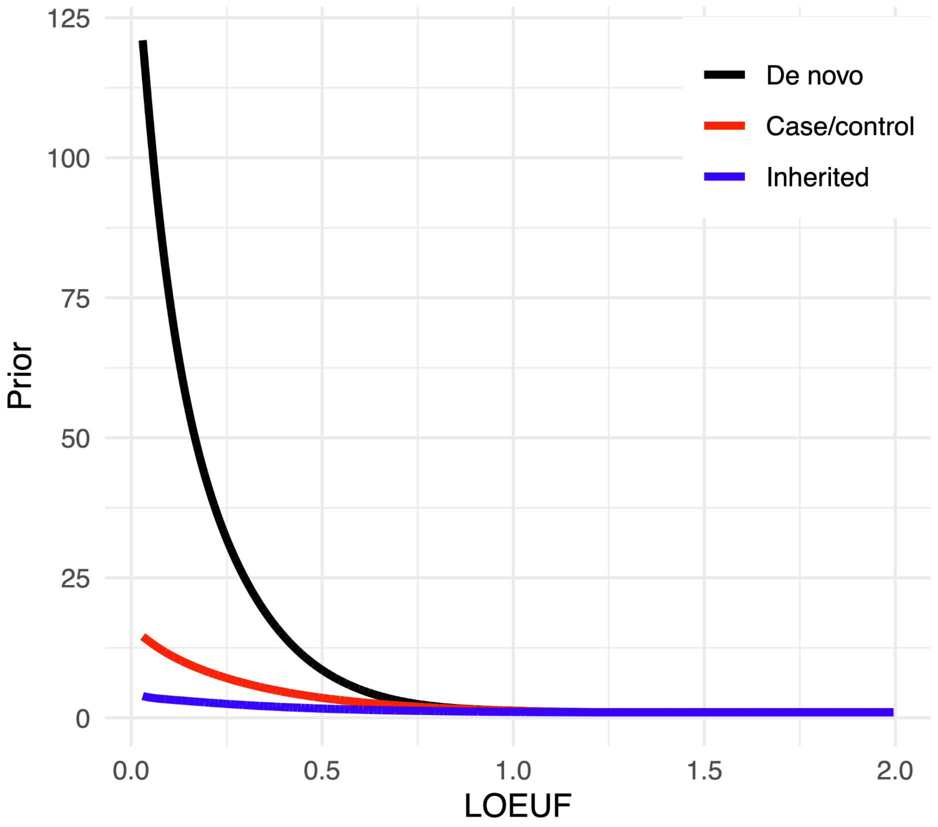


**Supplementary Figure 16**. TADA priors for predicted loss-of-function variants retrained using the DDD data. Priors are shown in black for the *de novo* analysis, red for case/control analysis, and blue for the transmitted/nontransmitted (“inherited”) analysis. For all lines, genes with lower LOEUF values (more constrained) have higher priors.

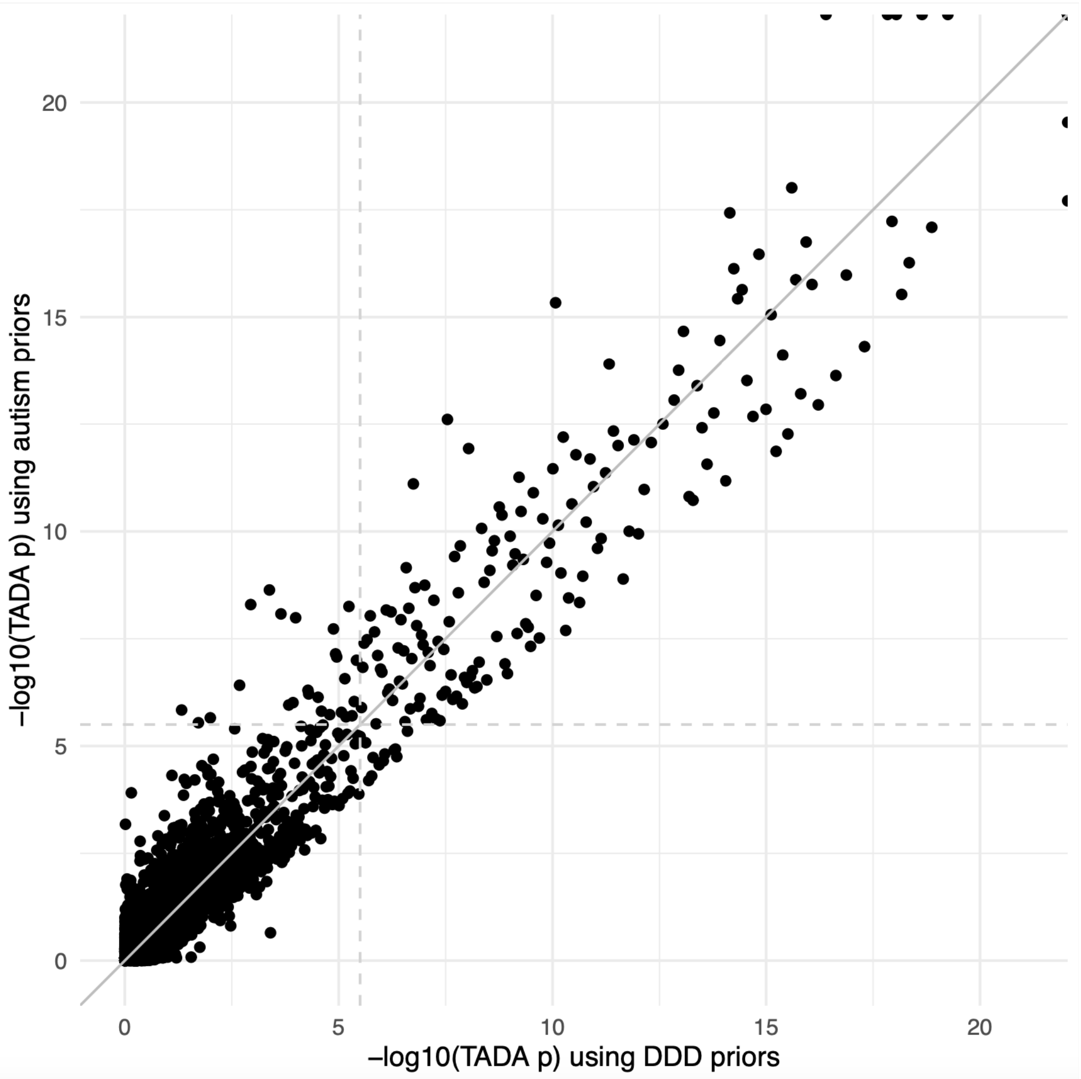


**Supplementary Figure 17**. Comparison of the TADA results when using mutation rates and priors from this study ("DDD priors", x) to those when using mutation rates and priors from the Fu et al. study of autism ("autism priors", y). The Pearson correlation coefficient is 0.833 between these two results. The solid gray line indicates y = x; the two dashed gray lines p ~$2.8 \times 10^{-6}$, the exome-wide significance threshold.

### Supplementary Tables

| | Mean (Median) Number of Rare Variants | | | |
|---|---|---|---|---|
| | Cases | Fathers | Mothers | Controls |
| **Synonymous** | 71.8 (71) | 70.5 (70) | 70.3 (70) | 72.0 (71) |
| **Missense** | 137.6 (137) | 134.4 (134) | 134.1 (134) | 137.6 (137) |
| **Inframe** | 2.98 (3) | 2.85 (3) | 2.86 (3) | 2.94 (3) |
| **Splice LoF** | 2.31 (2) | 2.23 (2) | 2.22 (2) | 2.21 (2) |
| **Stop gained** | 3.79 (4) | 3.63 (3) | 3.61 (3) | 3.63 (3) |
| **Frameshift** | 3.83 (4) | 3.66 (3) | 3.67 (3) | 3.65 (3) |

**Supplementary Table 1**. The mean (and median) number of rare (allele frequency < 0.001) variants in European genetic ancestry developmental disorder cases, their parents (split by sex), and controls, stratified by mutation consequence.

**Supplementary Table 2**. Results of case versus control regressions for six sets of DD cases compared to 3,943 ancestry-matched controls. We report the odds ratios from logistic regressions. The six sets of cases of inferred European genetic ancestry were: (1) all DD cases ("all", n=10,644), (2) all DD cases in complete trios ("all_trios", n=7,854), (3) all DD cases in non-trios ("non_trios", n=2,790), (4) trio cases with one or more affected parents ("affpar", n=1,246), (5) trio cases with a likely *de novo* diagnosis ("dndiag", n=2,679), and (6) trio cases with unaffected parents and no *de novo* diagnosis ("unaffpar_noDNdiag", n=4,183). Regressions were corrected for the first 20 principal components, sex, and the number of rare variants per person. Six mutation consequences are tested: predicted loss-of-function (lof), all missense (mis), all synonymous (syn), LOFTEE high-confidence lof (lof_HC), missense variants with CADD ≥20 (mis_CADD20), and missense variants with MPC ≥2 (mis_MPC2). Nine gene categories are tested: all genes ("all"), constrained genes ("cons", pLI ≥0.9), constrained monoallelic developmental disorder (DD)-associated genes ("cons_dd"), constrained genes that are not DD-associated ("cons_not_dd"), DD-associated genes ("dd"), genes that are not DD-associated ("not_dd"), the top decile of genes when scored by the missense observed/expected upper bound fraction ("top1moeuf"), unconstrained genes (pLI < 0.9, "uncons"), and unconstrained genes that are not DD-associated ("uncons_notdd").

**Supplementary Table 3**. Comparing the odds ratios from multiple case versus control regressions. We compared the results from (1) DD cases in non-trios ("non_trios", n=2,790), (2) trio cases with one or more affected parents ("affpar", n=1,246), and (3) trio cases with a likely *de novo* diagnosis ("dndiag", n=2,679) against those from trio cases with unaffected parents and no *de novo* diagnostic variant ("unaffpar_noDNdiag", n=4,183). To evaluate significant differences, we used a Wald test to obtain Z-scores that were transformed into p-values. We compared the results for LOFTEE high-confidence lof (lof_HC) and missense variants with CADD ≥ 20 (mis_CADD20) for four gene sets: all genes (“all”), constrained genes (“cons”, pLI ≥0.9), DD-associated genes (“dd”), and unconstrained genes that are not DD-associated (“uncons_notdd”).

**Supplementary Table 4**. Results of case versus control regressions for three sets of DD cases compared to 3,943 ancestry-matched controls when removing known *de novo* variants. We report the odds ratios from logistic regressions. The three sets of cases of inferred European genetic ancestry were: (1) trio cases with one or more affected parents (“affpar_noDNM”, n=1,246), (2) trio cases with a likely *de novo* diagnosis (“dndiag_noDNM”, n=2,679), and (3) trio cases with unaffected parents and no *de novo* diagnosis (“unaffpar_noDNdiag_noDNM”, n=4183). Regressions were corrected for the first 20 principal components, sex, and the number of rare variants per person. Six mutation consequences are tested: predicted loss-of-function (lof), all missense (mis), all synonymous (syn), LOFTEE high-confidence lof (lof_HC), missense variants with CADD ≥20 (mis_CADD20), and missense variants with MPC ≥2 (mis_MPC2). Nine gene categories are tested: all genes (“all”), constrained genes (“cons”, pLI ≥0.9), constrained monoallelic developmental disorder (DD)-associated genes (“cons_dd”), constrained genes that are not DD-associated (“cons_not_dd”), DD-associated genes (“dd”), genes that are not DD-associated (“not_dd”), the top decile of genes when scored by the missense observed/expected upper bound fraction (“top1moeuf”), unconstrained genes (pLI < 0.9, “uncons”), and unconstrained genes that are not DD-associated (“uncons_notdd”).

**Supplementary Table 5**. Results from the transmission disequilibrium test (TDT) comparing the number of transmitted, rare variants to nontransmitted rare variants for four sets of DD trios. In these analyses, we used DD cases in complete trios of all inferred genetic ancestry groups. If there were multiple affected children per family, one was selected as the representative sample. The four trio groups tested were: (1) all trios (“all”, n=9,305), (2) trios with one or more affected parents (“affpar”, n=1,306), (3) trios with a likely diagnostic *de novo* variant (“dndiag”, n=3,155), and (4) trios with unaffected parents and no diagnostic *de novo* variant (“unaffpar_noDNdiag”, n=5,124). Five mutation consequences are tested: predicted loss-of-function (lof), LOFTEE high-confidence lof (lof_HC), missense variants with CADD ≥20 (mis_CADD20), missense variants with MPC ≥2 (mis_MPC2), and all synonymous variants (synonymous). Nine gene sets are tested: all genes (“all”), constrained genes (“cons”, pLI ≥0.9), constrained monoallelic developmental disorder (DD)-associated genes (“cons_dd”), constrained genes that are not DD-associated (“cons_not_dd”), DD-associated genes (“dd”), genes that are not DD-associated (“not_dd”), the top decile of genes when scored by the missense observed/expected upper bound fraction (“top1moeuf”), unconstrained genes (pLI < 0.9, “uncons”), and unconstrained genes that are not DD-associated (“uncons_notdd”).

**Supplementary Table 6**. Comparing the transmission disequilibrium (TDT) test results from various DD case sets. We compared the results from (1) trio cases with one or more affected parents (“affpar”, n=1,306) and (2) trio cases with a likely *de novo* diagnosis (“dndiag”, n=3,155) against those from trio cases with unaffected parents and no *de novo* diagnostic variant (“unaffpar_noDNdiag”, n=5,124). All inferred genetic ancestries were used here, but only one affected child per family was selected. To evaluate significant differences, we used a $\chi^2$ test comparing the transmitted to nontransmitted variant counts to obtain a p-value. We compared the results for LOFTEE high-confidence lof (lof_HC) and missense variants with CADD ≥ 20 (mis_CADD20) for four gene sets: all genes (“all”), constrained genes (“cons”, pLI ≥0.9), DD-associated genes (“dd”), and unconstrained genes that are not DD-associated (“uncons_notdd”).

**Supplementary Table 7**. Comparing the burden of rare variants inherited from mothers versus that inherited from fathers in a set of 6,087 developmental disorder cases that are in complete trios, both parents are considered unaffected, and all three individuals in the family are of European genetic ancestry. For each combination of mutation consequence and gene category, the counts from mothers (“from_mom”) and from fathers (“from_dad”) are compared with a chi-square test (“p_value”). Six mutation consequences are tested: predicted loss-of-function (lof), all missense (mis), all synonymous (syn), LOFTEE high-confidence lof (lof_HC), missense variants with CADD ≥20 (mis_CADD20), and missense variants with MPC ≥2 (mis_MPC2). Nine gene categories are tested: all genes (“all”), constrained genes (“cons”, pLI ≥0.9), constrained monoallelic developmental disorder (DD)-associated genes (“cons_dd”), constrained genes that are not DD-associated (“cons_not_dd”), DD-associated genes (“dd”), genes that are not DD-associated (“not_dd”), the top decile of genes when scored by the missense observed/expected upper bound fraction (“top1moeuf”), unconstrained genes (pLI < 0.9, “uncons”), and unconstrained genes that are not DD-associated (“uncons_notdd”).

**Supplementary Table 8**. Results of a case versus case regression analysis comparing developmental disorder cases with (n=879) and without (n=3,309) environmental exposures (i.e., premature birth, maternal diabetes, and/or exposure to antiepileptic medications in utero). Here, we used only individuals of European genetic ancestry that were in complete trios, had unaffected parents, and did not have a *de novo* diagnosis. Regressions were corrected for the first 20 principal components, sex, and the number of rare variants per person. Six mutation consequences are tested: predicted loss-of-function (lof), all missense (mis), all synonymous (syn), LOFTEE high-confidence lof (lof_HC), missense variants with CADD ≥20 (mis_CADD20), and missense variants with MPC ≥2 (mis_MPC2). Nine gene categories are tested: all genes (“all”), constrained genes (“cons”, pLI ≥0.9), constrained monoallelic developmental disorder (DD)-associated genes (“cons_dd”), constrained genes that are not DD-associated (“cons_not_dd”), DD-associated genes (“dd”), genes that are not DD-associated (“not_dd”), the top decile of genes when scored by the missense observed/expected upper bound fraction (“top1moeuf”), unconstrained genes (pLI < 0.9, “uncons”), and unconstrained genes that are not DD-associated (“uncons_notdd”).

| Group | N | Effect size (95% CI) | P-value |
|---|---|---|---|
| All | 9305 | 0 (0 – 0) | 0.005 |
| European | 7037 | 0 (0 – 0) | 0.035 |
| Unaffected parents, no DNM diagnosis | 5124 | 0 (0 – 0) | 0.028 |
| DNM diagnosis | 3155 | 0 (0 – 0) | 0.714 |
| Affected parents | 1306 | 0.0005 (0.000 – 0.0119) | 0.012 |

**Supplementary Table 9**. Differences in the $s_{het}$ burden scores for developmental disorder (DD) cases compared to their parents. Shown are scores made when using rare missense variants with CADD ≥ 25 and MPC ≥ 2 for five sets of trios. In the case where a family had multiple affected children, only one child (and thereby complete trio) was used. For these analyses, all known *de novo* variants were removed from calculations. Bootstrapping with 1000 replicates was used to determine the median difference in $s_{het}$ burden scores as well as the 95% confidence intervals (CI). P-values are from the Wilcoxon rank sum test.

**Supplementary Table 10**. Results from the Transmission and De Novo Association (TADA) analysis for 17,646 autosomal genes tested. For every gene, we provide the gene symbol (“symbol”), Ensembl ENSG (“gene_id”), HGNC ID (“num_hgnc_id”), chromosome (“chrom”), mutation rates (“mut.Y”), LOEUF score, DDD-trained TADA priors (“prior.X.Y”), Bayes Factors (“BF_X_Y”), combined Bayes Factors for mutation consequences (“cBF_Y”), TADA q-value (“qval”), TADA p-value (“tada_p”), and variant counts. For mutation rates and variant counts, we provide these for pLoF variants (referred to as “ptv” to match the TADA code), missense variants with MPC ≥ 2 (“misB”), missense variants with 1 ≤ MPC < 2 (“misA”), all missense variants (“mis”), and synonymous variants (“syn”). For priors, Bayes Factors, and combined Bayes Factors, we provide this information for pLoF variants (“ptv”), missense variants with MPC ≥ 2 (“misB”), and missense variants with 1 ≤ MPC < 2 (“misA”) as these are the three mutation consequences included in the analysis. The priors, Bayes Factors, and variant counts are further split by their inheritance class: *de novo* (“dn”), case and control counts that are used in the case/control analysis (“case”, “control”, and “cc”), and transmitted to untransmitted variants in the inherited variant analysis (“in”, “inh”, “inh.t” for transmitted, and “inh.u” for untransmitted). As an example, the prior for a misB case/control variant would be “prior.cc.misb”.

**A) Variant type**

| | All significant (n=269) | Known significant (n=244) | New significant (n=25) | Not significant (n=17,377) |
|---|---|---|---|---|
| **pLoF** | 66.7 | 66.5 | 76.0 | 49.4 |
| **misB** | 27.1 | 27.2 | 21.6 | 19.1 |
| **misA** | 6.2 | 6.3 | 2.4 | 31.5 |

**B) Inheritance type**

| | All significant (n=269) | Known significant (n=244) | New significant (n=25) | Not significant (n=17,377) |
|---|---|---|---|---|
| ***De novo*** | 97.2 | 97.3 | 88.8 | 46.5 |
| **Case/control** | 2.5 | 2.4 | 9.7 | 39.6 |
| **Inherited** | 0.3 | 0.3 | 1.5 | 13.9 |

**Supplementary Table 11.** Percent Bayes Factor contributions from TADA split by variant (a) and inheritance (b) type. This is shown for all significant ($p < 2.8 \times 10^{-6}$) genes (n = 269), significant genes with some prior association to developmental disorders ("Known significant"; n = 244), significant genes with limited to no prior association to developmental ("New significant"; n = 25), and all other genes ("Not significant"; n = 17,377). Variant types are predicted loss-of-function (pLoF), missense variants with MPC ≥ 2 (misB), and missense variants with 1 ≤ MPC < 2 (misA).

**Supplementary Table 12.** Results of case versus control regressions for trios with a likely diagnostic *de novo* variant when keeping or removing those trios with one or more affected parents and when keeping or removing known *de novo* variants. The four regressions compared here are testing: (1) all trios with a diagnostic *de novo* variant for all rare variants ("dndiag", n=2,679), (2) trios with a diagnostic *de novo* variant and only unaffected parents for all rare variants ("dndiag_noAffectedParents"; n=2,525), (3) all trios with a diagnostic *de novo* variant for rare variants after removing known *de novo* variants ("dndiag_noDNM"), (4) trios with a diagnostic *de novo* variant and only unaffected parents for rare variants after removing known *de novo* variants ("dndiag_noDNM_noAffectedParents"). In all regressions, the cases were compared to 3,943 controls of inferred European genetic ancestry. Regressions were corrected for the first 20 principal components, sex, and the number of rare variants per person. Three mutation consequences are tested: LOFTEE high-confidence lof (lof_HC), missense variants with CADD ≥ 20 (mis_CADD20), and all synonymous (syn). Four gene sets were tested: all genes ("all"), constrained genes ("cons", pLI ≥0.9), DD-associated genes ("dd"), and unconstrained genes that are not DD-associated ("uncons_notdd").

**A) Predicted loss-of-function (pLoF) variants**

| | **DDD** | **Autism** |
|---|---:|---:|
| ***De novo* pLoF** | 120.98 | 17.16 |
| **Case/control pLoF** | 14.52 | 3.10 |
| **Inherited pLoF** | 3.90 | 1.56 |

**B) Missense variants**

| | **DDD** | **Autism** |
|---|---:|---:|
| ***De novo* misB** | 19.82 | 14.15 |
| **Case/control misB** | 2.96 | 3.08 |
| **Inherited misB** | 1.02 | 1.59 |
| ***De novo* misA** | 3.42 | 1.68 |
| **Case/control misA** | 1.43 | 1.05 |
| **Inherited misA** | 1.02 | 1.05 |

**Supplementary Table 13.** Comparison of priors used in TADA between the Fu et al. autism paper and those used here that were retrained using DDD data. In A) are the maximum priors for predicted loss-of-function (pLoF) variants. In B) are the priors for missense variants, split into "misA" (1 ≤ MPC < 2 ) and "misB" (MPC ≥ 2) categories as in the Fu et al. paper. For both tables, variants are categorized by their inheritance type: *de novo*, case/control, and inherited.
